# Prediction of the infecting organism in peritoneal dialysis patients with acute peritonitis using interpretable Tsetlin Machines

**DOI:** 10.1101/2025.03.03.25323096

**Authors:** Olga Tarasyuk, Anatoliy Gorbenko, Matthias Eberl, Nicholas Topley, Jingjing Zhang, Rishad Shafik, Alex Yakovlev

## Abstract

**Motivation:** The analysis of complex biomedical datasets is becoming central to understanding disease mechanisms, aiding risk stratification and guiding patient management. However, the utility of computational methods is often constrained by their lack of interpretability and accessibility for non-experts, which is particularly relevant in clinically critical areas where rapid initiation of targeted therapies is key.

**Results:** To define diagnostically relevant immune signatures in peritoneal dialysis patients presenting with acute peritonitis, we analysed a comprehensive array of cellular and soluble parameters in cloudy peritoneal effluents. Utilising Tsetlin Machines (TMs), a logic-based machine learning approach, we identified pathogen-specific immune fingerprints for different bacterial groups, each characterised by unique biomarker combinations. Unlike traditional ‘black box’ machine learning models such as artificial neural networks, TMs identified clear, logical rules in the dataset that pointed towards distinctly nuanced immune responses to different types of bacterial infection. This demonstrates unambiguously that even when infecting the same anatomical location and causing clinically indistinguishable symptoms, each type of pathogens interacts in a specific way with the body’s immune system. Importantly, these immune signatures could be easily visualised to facilitate their interpretation, thereby not only enhancing diagnostic accuracy but also potentially allowing for rapid, accurate and transparent decision-making based on the patient’s immune profile. This unique diagnostic capacity of TMs could help deliver clear and actionable insights such as early patient risk stratification and support early and informed treatment choices in advance of conventional microbiological culture results, thus guiding antibiotic stewardship and contributing to improved patient outcomes.

**Availability and implementation:** All underlying tools for the present analysis are available at https://github.com/anatoliy-gorbenko/biomarkers-visualization. The anonymised patient data underlying this article will be shared on reasonable request to the corresponding authors.

## 1. Introduction

Reliable, rapid and accurate diagnosis of infection remains an unmet clinical need. Microbiological culture can take several days to generate results and is often impacted by inadequate sample quality, contamination and problems with fastidious or slow-growing organisms (Chakera et al., 2018). Molecular techniques to detect pathogens such as mass spectrometry or polymerase chain reaction equally depend on sample quality, may yield negative results at low pathogen numbers and do not discriminate between live and dead organisms. Finally, host biomarkers in patients presenting with suspected infections are often relatively unspecific due to the highly dynamic and individual nature of the early immune response. Despite promising advances, no single biomarker is sufficiently specific or sensitive to accurately predict the presence of an infection or indeed the type or even species of causative pathogen (Chakera et al., 2018). Factors such as patient age and gender, comorbidities, infection severity, pathogen type and virulence can all influence biomarker expression patterns, as can medication. This creates a challenge for the accurate identification of robust immune signatures that could guide more rapid diagnosis and targeted antibiotic treatment (Aufricht et al., 2017). The complexity of biomarker profiles necessitates sophisticated multi-parameter analysis techniques, leading to an increased interest in applying machine learning (ML) methods to biomedical datasets, aiming to improve patient stratification and tailor therapies more effectively.

ML models such as support vector machines, artificial neural networks (ANNs) and random forests have been successfully applied to biomedical datasets (Ahsan et al., 2022; Peiffer-Smadja et al., 2020), including those from our own work in patients with urinary tract infection (Gadalla et al., 2019), peritoneal dialysis (PD)-related peritonitis (Zhang et al., 2017) and sepsis (Burton et al., 2024). However, the lack of explainability in most ML models – the lack of understanding and hence of confidence in what they show – is a major barrier to their wider clinical adoption. Many ML methods, especially intricate ones like ANNs, often function as ‘black boxes’ (Rudin, 2019), making it difficult for scientists, healthcare professionals and regulatory authorities to understand and trust the basis of ML-based predictions.

New ML techniques that are better interpretable are being developed to bridge this gap, such as approaches using decision trees (Mienye & Jere, 2024), probabilistic and fuzzy logic (Zheng et al., 2024), or focusing on explainable artificial intelligence tools and post-hoc interpretability methods like SHapley Additive exPlanations (SHAP) (Burton et al., 2024; Salih et al., 2024). However, their computational demands, difficulty of standardisation and challenges in usability continue to hinder their widespread adoption. Indeed, current explainable artificial intelligence tools often provide explanations in technical or abstract terms that may not align with the clinical reasoning process. The creation of user-friendly interfaces and clinical decision-support systems that integrate seamlessly with existing workflows remains an area of active research.

This paper aims at providing explainability in decision making via employing a relatively new logic-based ML algorithm called Tsetlin Machine (TM), for which explainability is an intrinsic feature. TMs rely on the collective behaviour of learning automata (Narendra & Thathachar, 2012; Tsetlin, 1973; Varshavsky & Pospelov, 1988). As part of their training and inference, TMs generate a set of conjunctive logical statements (logical clauses that can be viewed as directly interpretable inference rules), which vote for or against each class, thus justifying the decision making. TMs have considerably fewer hyperparameters to tune than other ML methods. These are highly interpretable since their model prediction is carried out via proposition logic clauses. The logical rules can be naturally visualised, which further eases their understanding and interpretation by specialists. The fact that TMs use Boolean (*i.e.* semi-quantitative) features as input data, unlike other ML methods that operate with continuous numerical values, makes them particularly attractive for deciphering biological processes. As a consequence, for clinical use, they may allow easier translation of complex datasets generated using laborious techniques into the design of simpler methods such as lateral flow tests, while maintaining competitive accuracy.

We describe the application of TM-based analysis techniques to a set of soluble and cellular biomarkers measured previously in individuals presenting with acute PD-related peritonitis (Zhang et al., 2017). PD is a life-saving renal replacement therapy used to manage end-stage kidney disease by removing waste products, excess fluid and toxins from the body, utilising the peritoneal membrane lining the abdominal cavity as a semipermeable filter. Despite its effectiveness and convenience for many patients, PD carries the risk of peritonitis, a severe infection of the peritoneal cavity (Li et al., 2022). Peritonitis is a significant complication that can arise from contamination during catheter handling, bowel leakage and other reasons, and is associated with significant morbidity, treatment failure and in some cases death (Cho et al., 2024). Moreover, any inflammatory episode of peritonitis may cause scarring and thickening of the peritoneal membrane (Fielding et al., 2014) and potentially contribute to treatment failure. Timely diagnosis and antimicrobial intervention are thus key to successful treatment (Chakera et al., 2018). However, despite the recognition more than three decades ago that levels of inflammatory markers are elevated in the peritoneal effluent hours prior to the manifestation of overt clinical symptoms (Betjes et al., 1996), a simple biomarker-based lateral flow test for early peritonitis was only recently developed (Goodlad et al., 2020; Htay et al., 2024) and has not been widely adopted.

It is well recognised that different classes (Gram-positive and Gram-negative bacteria) and species of micro-organisms result in different patient outcomes and that infections with them give rise to distinct sets of biomarkers (‘immune fingerprints’) (Liuzzi et al., 2016; Zhang et al., 2017). Immune fingerprints have shown promise for rapid point of care prediction of infection (Gadalla et al., 2019) and causative pathogen (Burton et al., 2024; Zhang et al., 2017) in other infectious contexts, potentially allowing early risk stratification and targeted antibiotic treatment. By combining biomarker measurements during acute peritonitis and logic-based inference approaches as offered by TMs, we now demonstrate the power of interpretable ML models to analyse complex biomarker signatures. Our findings may have immediate diagnostic implications, potentially guiding appropriate antibiotic treatment before conventional microbiological culture results become available.

## 2. Materials and Methods

### 2.1 Patients

The study cohort comprised 82 adults receiving peritoneal dialysis (PD) who were admitted between 2008 and 2016 to the University Hospital of Wales in Cardiff (UK) with acute peritonitis. Clinical diagnosis of peritonitis was based on the presence of abdominal pain and cloudy peritoneal effluent with >100 white blood cells per mm^3^ (Li et al., 2022). According to the microbiological analysis of the effluent, peritonitis episodes were defined as culture-negative or as confirmed bacterial infections by a Gram-positive or Gram-negative organism (**Supplemental Tables S1 and S2**). Cases of fungal infection and mixed or unclear culture results were excluded. The study was approved by the South East Wales Local Ethics Committee (04WSE04/27) and registered on the UK Clinical Research Network Study Portfolio under reference number #11838 “Patient immune responses to infection in peritoneal dialysis” (PERIT-PD). All individuals provided written informed consent.

### 2.2 Machine learning with interpretable Tsetlin Machine

TMs leverage the collective behaviour of learning automata and bases its inference on interpretable logic-based rules, specifically conjunctive clauses (Granmo, 2018; Lei et al., 2020). These clauses logically link together input features, thereby creating distinct patterns that represent different classes, enabling TMs to make transparent and interpretable predictions of infecting organisms in peritoneal dialysis patients with acute peritonitis. TMs are an actively evolving field of research where novel architectures and training methods are being developed and becoming available via the GitHub repository (Centre for Artificial Intelligence Research, 2018). Here, the Python implementation of the basic *MulticlassTsetlinMachine* from the *pyTsetlinMachineParallel* package was used. The source code is available at https://github.com/cair/pyTsetlinMachineParallel.

A TM has three major hyperparameters affecting its performance and defining a balance between clauses generalisation and specialisation, namely: the number of clauses per class *C* (*i.e.* the number of logical rules used for inference), the voting threshold *T* and the learning sensitivity *s*. During the tuning of TM hyperparameters, the number of clauses *C* was set to 20 in the present study, with ten *positive* clauses creating class patterns and ten *negative* clauses generating patterns for the counter-class(es). This provided a reasonable balance between achieving high classification accuracy and maintaining ease of comprehension and interpretation of the resulting logical rules. The voting threshold *T* and the learning sensitivity *s* were set to their optimum values as described (Tarasyuk, Rahman, et al., 2023). In particular, the global optimum of the voting threshold *T* for the given number of clauses *C* approximates to the square root of *C*/2, which maximises voting power of each clause according to Jagiellonian compromise for qualified majority in the Penrose’s square root voting system (Penrose, 1946). The optimal value of *s* scales as the logarithm of *C* and was chosen experimentally at each classification step (**Figure 1**).

**Figure 1.**
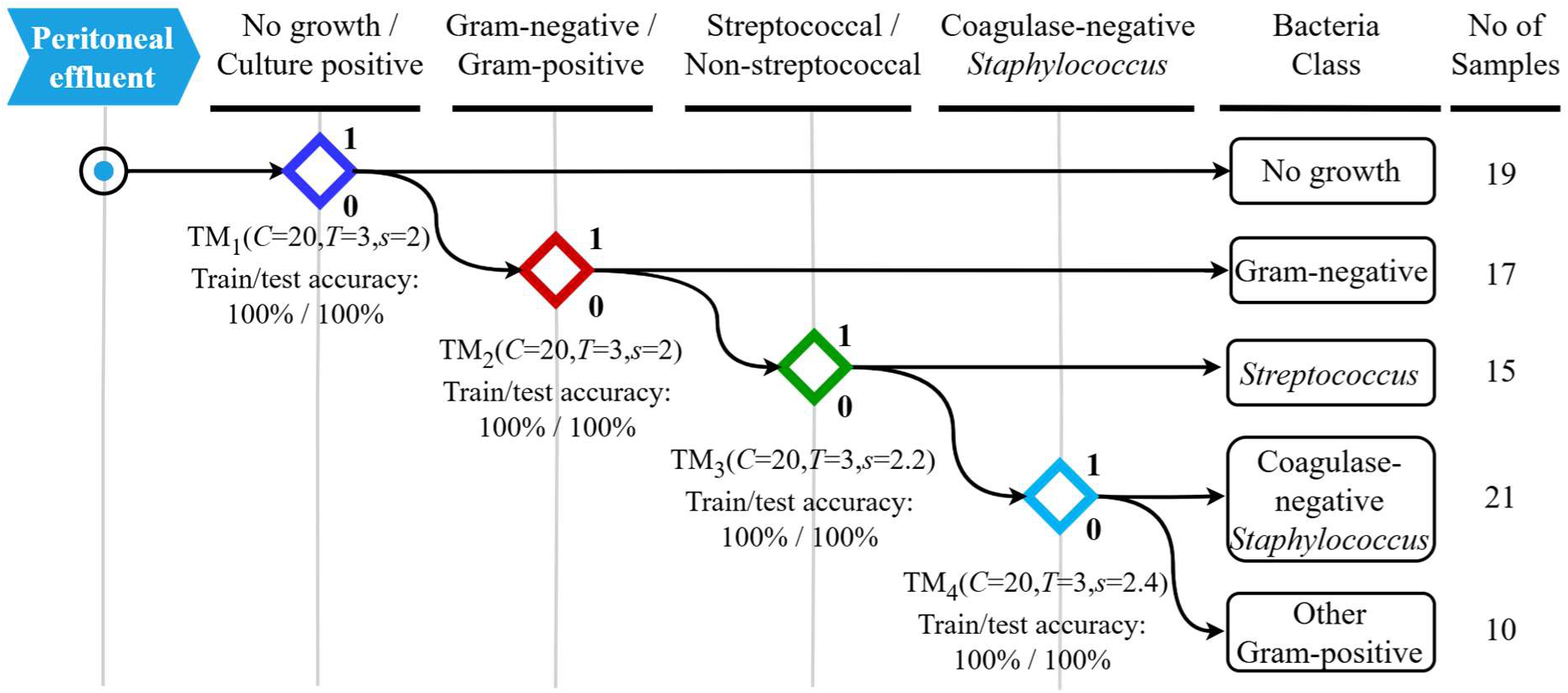
Hierarchical classification methodology to identify local immune fingerprints associated with peritonitis caused by different types of bacteria. A binary decision tree attempts to predict the causative organism in the following order: 1) discrimination between episodes of peritonitis that yielded no microbiological growth versus culture-positive episodes; 2) discrimination between episodes caused by Gram-negative bacteria within the culture-positive group of patients; 3) discrimination between episodes caused by streptococcal organisms versus episodes caused by non-streptococcal Gram-positive bacteria; 4) discrimination between episodes caused by coagulase-negative *Staphylococcus* versus episodes caused by other Gram-positive bacteria.

Tsetlin automata (Narendra and Thathachar, 2012) in the TM serve as fundamental units for decision-making and learning, similar to artificial neurons in ANNs, although their roles and mechanisms differ significantly (Lei et al., 2020; Tarasyuk et al., 2023a). The ANNs used in our previous study (Zhang et al., 2017) were adopted here as reference ML model, configured with 40,000 neurons in the hidden layer for multi-class classification and 13,000 neurons for binary classification. This setup ensured comparable complexity to TMs configured with 20 clauses per class, utilising up to 39,200 and 12,800 Tsetlin automata at maximum configuration, respectively.

### 2.3 Data preprocessing

#### 2.3.1 Data imputation

The original dataset had some missing biomarker values due to incomplete or failed measurements (**Supplemental Table S4**) (Zhang et al., 2017). Missing data were imputed to fit gaps by adopting Multivariate Imputation by Chained Equations (MICE) implemented in R (van Buuren & Groothuis-Oudshoorn, 2011), which imputes an incomplete feature by generating synthetic values considering their relationship with other biomarkers.

#### 2.3.2 Data Booleanisation

TMs operate on Boolean input data where each input feature can take only one of two values, 1 (*i.e. True*) or 0 (*i.e. False*). To Booleanise the input dataset and convert biomarker values from quantitative to semi-quantitative Boolean features we used a simple binning method. For each biomarker we determined its range as the difference between the measured maximum and minimum values, which was then divided into equal intervals (bins). These intervals were then encoded using ‘one-hot encoding’ method, so that each biomarker is represented by a unique binary vector with the same length as the number of intervals. In this vector, only one element is set to 1 (*True*), indicating the presence of the biomarker value in that specific interval, and all other elements are set to 0 (*False*). For instance, using four semi-quantitative intervals per value, each of the 82 patient samples comprising 49 biomarkers was represented by 196 Boolean features (four bits/Boolean features per biomarker) (**Supplemental Figure S1**).

Booleanisation simplifies datasets by reducing their granularity, making it easier to analyse and interpret in certain applications, such as rule-based machine learning or visualisation. TM complexity, measured by the number of Tsetlin automata utilised, is directly proportional to the number of Boolean features. Each feature is linked via a dedicated Tsetlin automaton to each logical clause. Depending on the state of this automaton after the completion of training, a particular feature may either be included in or excluded from the certain logical clauses. Here, we experimented with two, three and four semi-quantitative intervals, revealing a trade-off between the complexity of the TM and its classification accuracy.

#### 2.3.3 Data balancing

The peritonitis cohort contained an unequal distribution of patients across different classes (**Figure 1**), which resulted in an unbalanced dataset. This usually leads to biasing ML models toward the majority class(es) and poor performance on the minority class(es). It is also essential for TMs to support even clauses development by ensuring that TM learning automata of different classes receive equal reinforcements. Therefore, at each stage of the hierarchical classification we first divided the Booleanised dataset into train and test subsets with an 80% to 20% split in a stratified fashion using the *train_test_split* function from the *sklearn* Python package. This ensured that relative class frequences were preserved in training and validation subsets. Second, to address class imbalance within the training subset, a random over-sampling technique was applied to the minority classes by selecting samples at random with replacement (this ensures the constant probability of selecting any specific sample) by employing *RandomOverSampler* from the *imblearn* Python package. This approach provided a balanced training dataset, facilitating equal reinforcement opportunities across classes.

## 3. Results

### 3.1 Hierarchical Classification

To identify organism-specific immune fingerprints and predict the causative pathogen, a wide range of cellular and soluble immune biomarkers was measured in the cloudy effluent of PD patients presenting with acute peritonitis (**Supplemental Tables S1 and S2)**. These biomarkers included frequencies and total numbers of infiltrating leukocyte populations as well as levels of inflammatory mediators and tissue damage-associated molecules, covering the breadth and complexity of the local immune response to infection (Zhang et al., 2017) (**Supplemental Table S3)**.

An initial attempt to define immune fingerprints that would simultaneously discriminate patients with all major groups of infecting organisms showed relatively low validation performance for different ML techniques such as support vector machines, random forests and artificial neural networks, despite high training accuracy (Zhang et al., 2017). Such overfitting could be explained by an insufficient size of the training dataset and complex relationships between different biomarkers. Reassuringly, TMs demonstrated considerable improvement in multiclass classification and allowed to reach 65% of test accuracy while maintaining 100% train accuracy on the whole dataset. For comparison, ANNs of a comparative complexity with 40,000 hidden nodes achieved a peak test accuracy of 53% (**Table 1**).

**Table 1.**
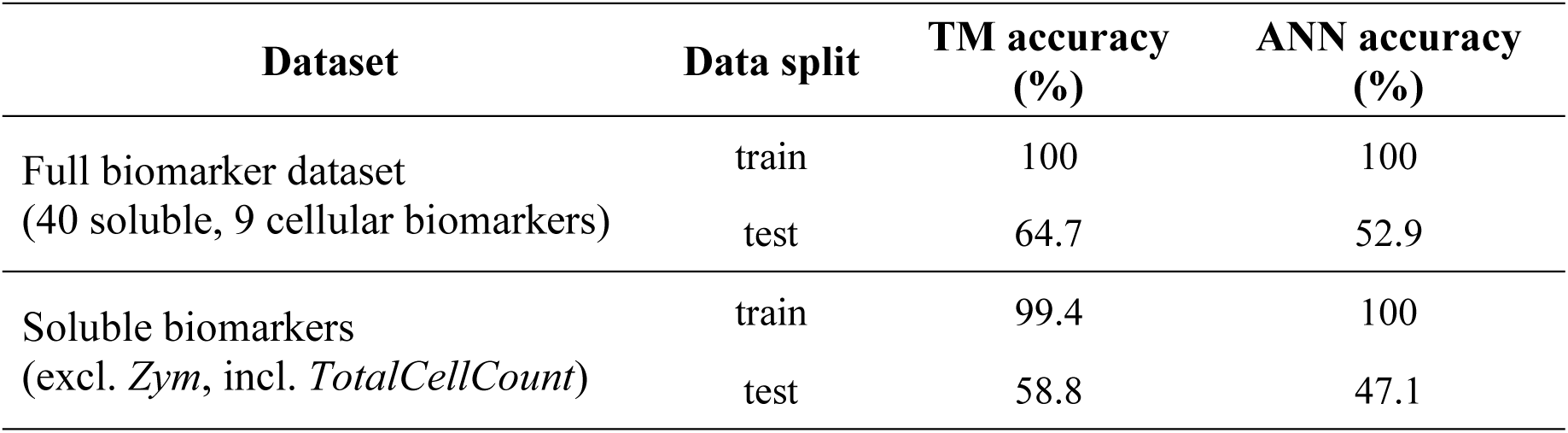
Multi-class classification train and test accuracy of TMs. The table shows the accuracy of a single TM trained to discriminate between five classes (No growth, Gram-negative, streptococcal, coagulase-negative *Staphylococcus*, and other Gram-positive bacteria) at once, using the whole biomarkers dataset and a subset of soluble biomarkers (excluding *Zym* and including *TotalCellCount*). TM accuracy was determined for 20 clauses across 4 ranges and a total of 39,200 learning automata. The rightmost column shows the peak accuracy achieved by an artificial neural network (ANN) of comparable complexity using 40,000 neurons, which is used as a benchmark value.

Whilst providing encouraging proof of concept for the validity of a TM based approach, such an accuracy would not be satisfactory for clinical use. Thus, we adopted a binary classification approach that focussed on discriminating between a certain class of bacterial infection and other cases of peritonitis, and which had already returned promising results for support vector machines (Zhang et al., 2017). We improved it further by structuring it in a hierarchical stepwise manner, resembling a binary decision tree attempting to predict the causative organism in the following order (**Figure 1**):

1. Discrimination between episodes of peritonitis that yielded no microbiological culture result (‘no growth’; *n*=19) and episodes where a bacterial pathogen was identified (‘culture-positive’; *n*=63);
2. Within the culture-positive group, discrimination between episodes caused by Gram-negative bacteria (*Acinetobacter baumannii*, *Enterobacter* spp., *Escherichia coli*, *Morganella morganii*, *Proteus vulgaris*, *Pseudomonas aeruginosa* and others; *n*=17) and episodes caused by Gram-positive bacteria (*n*=46);
3. Within the confirmed infections caused by Gram-positive bacteria, discrimination between episodes caused by streptococcal organisms (*Streptococcus* spp. and *Enterococcus* spp.; *n*=15) and episodes caused by other, non-streptococcal Gram-positive bacteria (*n*=31);
4. Within the confirmed infections caused by non-streptococcal Gram-positive bacteria, discrimination between episodes caused by coagulase-negative *Staphylococcus* (CNS; *n*=21) and episodes caused by other Gram-positive bacteria (*Staphylococcus aureus*, *Corynebacterium* spp. and others; *n*=10).

Employing specialised TMs at each step of this hierarchical classification process allowed to achieve remarkable 100% accuracy on both train and test datasets, with only twenty logical clauses (rules) used to support decision-making in favour of one or another type of bacterial infection. Three quantitative ranges per biomarker at each classification step were usually sufficient to reliably distinguish between different types of bacterial infection; using only two ranges failed to ensure 100% train and test accuracy, while employing more than four ranges added unnecessary redundancy and data/computational overheads (**Table 2**). As before, TMs consistently outperformed ANNs.

**Table 2.** Train and test accuracy of TMs at different stages of the hierarchical binary classification. The table shows how TM accuracy (with 20 clauses) depends on the number of semi-quantitative intervals used to quantise values of soluble biomarkers (excluding *Zym* and including *TotalCellCount*). The rightmost column reports the peak accuracy achieved by an artificial neural network (ANN) of comparable complexity (13,000 neurons), which is used as a benchmark value. TAs, Tsetlin automata.

| # | Classification step | Data split | TM accuracy (%) |  |  | ANN accuracy (%) |
| --- | --- | --- | --- | --- | --- | --- |
| | | | 2 ranges<br>(6,400 TAs) | 3 ranges<br>(9,600 TAs) | $\geq 4$ ranges<br>(12,800 TAs) | |
| 1 | No growth<br>vs Culture-positive cases | train | 98.0 | 100 | 100 | 100 |
|  |  | test | 94.1 | 100 | 100 | 82.4 |
| 2 | Gram-positive<br>vs Gram-negative bacteria | train | 98.6 | 100 | 100 | 100 |
|  |  | test | 92.3 | 100 | 100 | 76.9 |
| 3 | Gram-pos. streptococcal<br>vs non-streptococcal bacteria | train | 100 | 100 | 100 | 100 |
|  |  | test | 90 | 100 | 100 | 80 |
| 4 | Coag.-neg. *Staphylococcus*<br>vs other Gram-pos. bacteria | train | 100 | 100 | 100 | 100 |
|  |  | test | 100 | 100 | 100 | 85.7 |

### 3.2 Interpretability and Visualisation of Tsetlin Machine Clauses

The major advantage of TMs as compared to other ML algorithms is their natural interpretability and explainability. TMs make predictions based on a set of logical rules (clauses) generated during training, which explain the decision making and can be verified by specialists. A fragment of such rules created by the TMs to predict the presence of Gram-negative bacteria in patients presenting with acute peritonitis is shown as example in **Supplemental Figure S2**. These rules are represented in the form of conjunctive statements that include specific input features or their negations, thus creating a set of persistent sub-patterns (immune fingerprints) of the target class.

Although these rules were machine readable and easily interpretable, in their raw form they might still be difficult for humans to comprehend and explain, especially if the number of clauses and Boolean features involved was large. To solve this issue, we proposed a clause visualisation framework, which represented each clause as a biomarker-wise mask or stencil (**Supplemental Figure S3**). Each row of this stencil corresponded to a certain biomarker, while each biomarker was represented by a group of bits/pixels corresponding to different value ranges or concentration levels. These ranges were identified for each biomarker during the Booleanisation step as part of data pre-processing. A blue pixel meant that the biomarker value *must* be within that specific range to match the clause rule. Red meant that the biomarker value *must not* be in that range. Finally, white meant ‘ambivalent’, *i.e.* the biomarker value *may or may not* be in that range. An individual clause could thus be seen as a class template generalising certain common features of class samples from the training dataset. Each clause formulated a rule by identifying general patterns shared among a subset of patient samples within the same class, for which the clause would output *True* (**Supplemental Figure S3**). Collectively, the team of TM clauses determined the type of bacterial infection by evaluating the number of clauses that supported each classification hypothesis.

### 3.3 Tsetlin Machine Inference and Decision-Making

During TM inference, the input data sample was matched against all clauses of all classes. If a data sample perfectly aligned with the clause stencil, the clause would output *True*, indicating that the clause cast a vote suggesting the sample belonged to the designated class. Otherwise, the clause would output *False*, which means it abstained from voting. The class with the maximum sum of clause votes was returned as TM prediction. In this sense, TM inference based on clauses voting for and against each class fostered collaborative decision-making and ensured a thorough and holistic assessment. As example, **Figures 2 and 3** illustrate the inference process that enabled differentiation between Gram-negative and Gram-positive bacterial infections.

**Figure 2.**
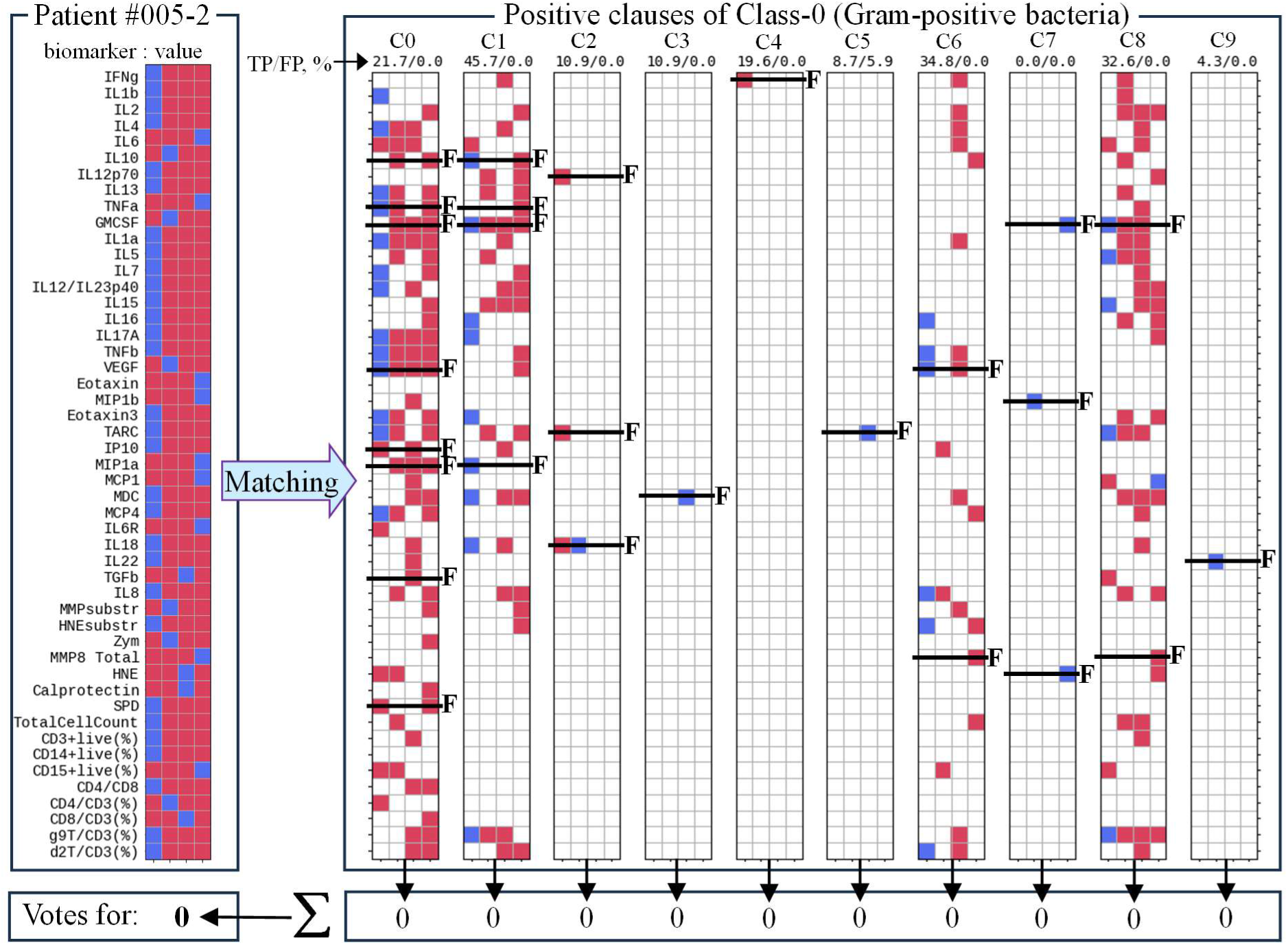
Example of TM inference supporting decision making in favour of Gram-positive bacterial infection. The figure shows the inference process using TM_2_ clauses trained to recognise Gram-positive (Class-0) bacterial infection by matching the patient sample against clause stencils followed by clause output summation and voting. Here, none of the clauses supported the hypothesis. *F*, biomarkers whose values lay outside the target range specified by the clauses, indicating the corresponding conjunct was *False*. Accuracy labels above each clause show the percentage of True Positive (TP) and False Positive (FP) predictions of individual clauses.

**Figure 3.**
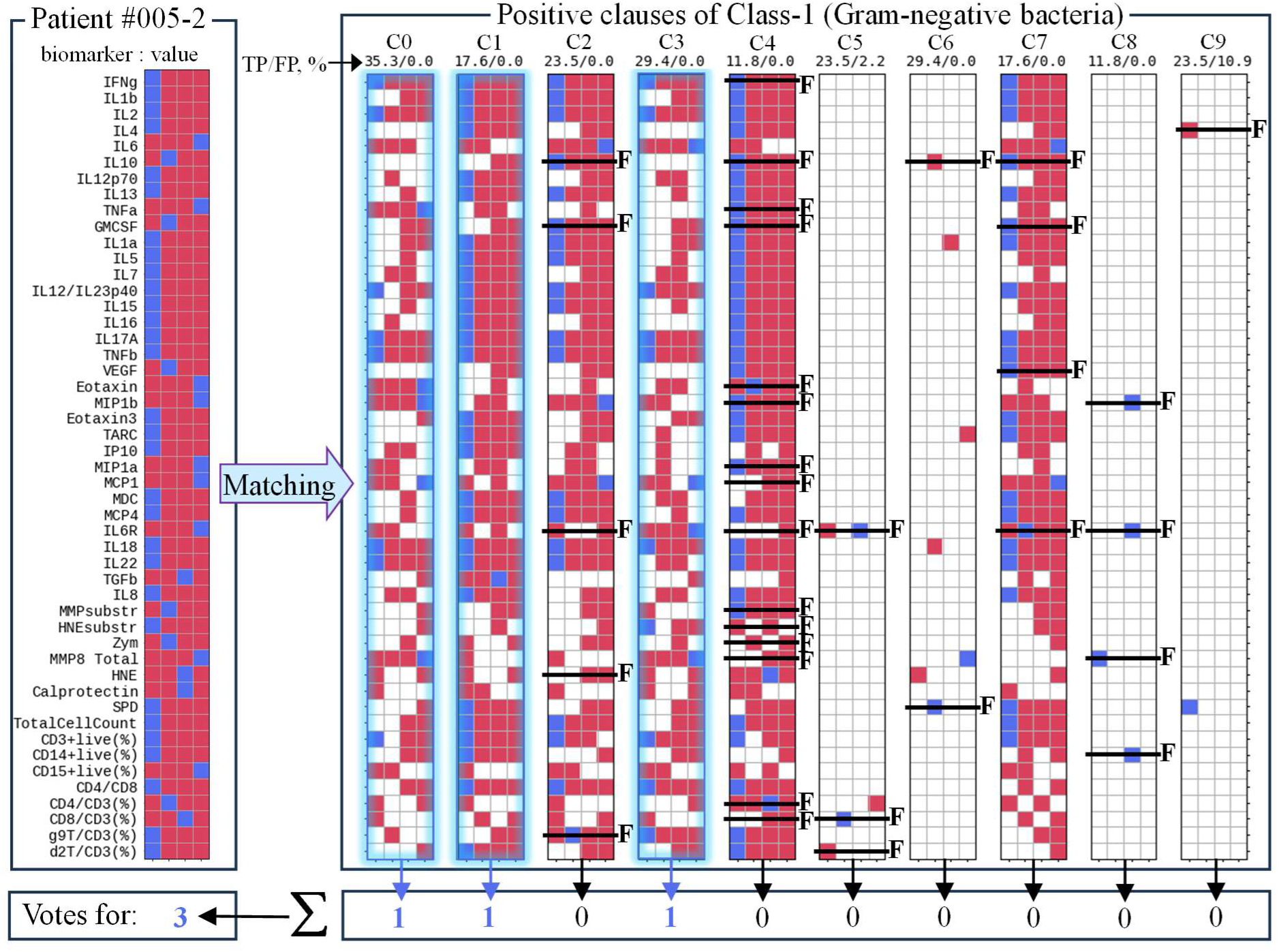
Example of TM inference supporting decision making in favour of Gram-negative bacteria infection. The figure shows the inference process using TM_2_ clauses trained to recognise Gram-negative (Class-1) bacteria by matching the patient sample against clause stencils followed by clause output summation and voting. Here, three out of ten clauses supported the hypothesis. *F*, biomarkers whose values lay outside the target range specified by the clauses, indicating the corresponding conjunct was *False*. Accuracy labels under each clause show the percentage of True Positive (TP) and False Positive (FP) predictions of individual clauses.

**Figure 2** shows how the data sample from patient 005-2 was matched against ten positive TM clauses trained to recognise and vote for Class-0 (Gram-positive bacteria). In this particular example, there was no match with any of these ten positive clauses. Biomarkers whose values lay outside the target range specified by the clauses were labelled ‘*F*’, indicating that the corresponding conjunct was *False*. Consequently, all ten clauses abstained from supporting the decision that patient 005-2 was infected with Gram-positive bacteria. The results of matching the same patient sample against positive clauses of Class-1 (Gram-negative bacteria) are shown in **Figure 3**. The data sample matched three clauses (C0, C1 and C3) out of ten. Ultimately, with three votes to zero, the patient 005-2 was classified as having a Gram-negative bacterial infection. Voting margin could be seen as an additional measure of confidence offered by TMs in decision making. TMs thus offered a useful mechanism of logical clauses which justified decision-making and could be easily interpreted, visualised and verified.

### 3.4 Identification of Key Biomarkers and Clause Minimisation

TMs facilitate the identification and ranking of key features (in this case, biomarkers) based on their impact on decision-making, whilst preserving logical relations between them. The initial step involves calculating the frequency with which each feature appears in the positive clauses of each class and negative clauses of the opposing class(es). Features that appear in clauses of both classes are the least significant for distinguishing them. The most significant are those that are unique to each class.

Here, the importance of each feature (a feature rank) was calculated by taking the absolute difference between the sum of occurrences of the feature in logical clauses that predicted the target class, and the sum of appearance of the feature in clauses in clauses that predicted the opposing class. A higher feature rank indicated that the feature had a strong association with one class over the other, making it highly influential in distinguishing between the two classes. Since each feature *x_i_* encoded a certain semi-quantitative interval for a particular biomarker, once the key features were identified, they were mapped back to their corresponding biomarkers. In the final step, the least significant biomarkers were removed from TM clauses one by one until the accuracy remained higher than the target threshold.

**Figure 4** presents the minimal sets of all biomarkers that achieved prediction accuracies of 90%, 95% and 100%, respectively, at different classification steps where each biomarker value was encoded using four semi-quantitative intervals. Perhaps not surprisingly, achieving higher accuracy required more biomarkers to be taken into consideration. For example, distinguishing between culture-positive infections and cases of no microbiological growth could be done with 90% accuracy using only 11 biomarkers. Achieving 95% required 13 biomarkers, while considering 21 biomarkers allowed 100% accuracy. At the same time, the later stages of hierarchical binary classifications required progressively fewer biomarkers. Taken together, these findings demonstrated that each type of microbiologically confirmed infection was associated with a distinct set of biomarkers that set it apart from infections with other organisms. For instance, accurate prediction of culture-positive episodes of peritonitis required cellular parameters such as the total cell count (*TotalCellCount*), the proportion of neutrophils amongst infiltrating immune cells (*CD15+live(%)*) and the proportion of Vδ2^+^ T cells amongst T cells (*d2T/CD3*), as well as a distinct set of cytokines and chemokines (**Figure 4**).

**Figure 4.**
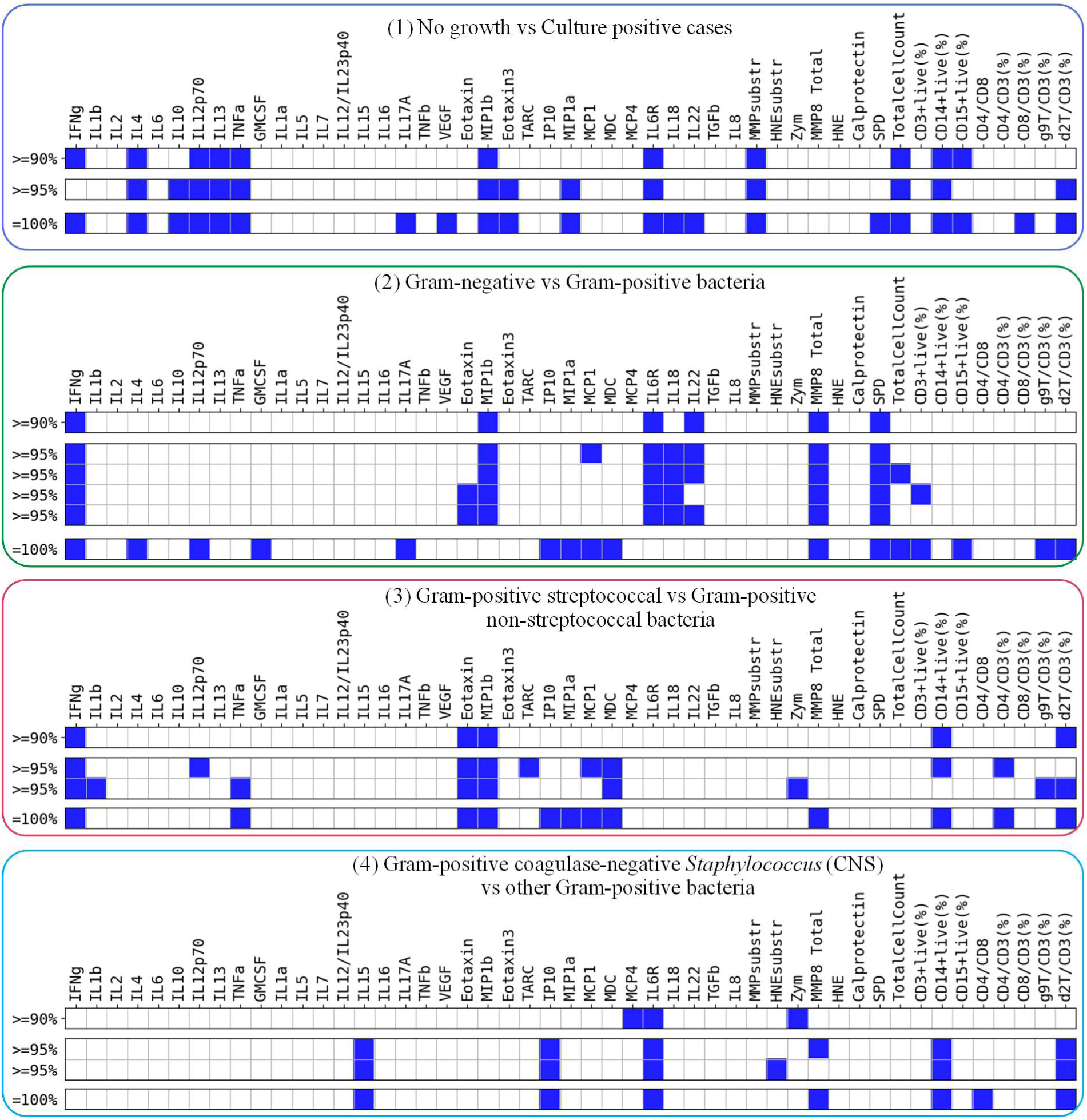
Minimised sets of soluble and cellular immune biomarkers for the case where each biomarker value was Booleanised by four semi-quantitative ranges. Figure shows the minimised set of soluble and cellular immune biomarkers needed to make predictions at different classification stages with the target accuracies of 90%, 95% and 100%, respectively.

### 3.5 Focus on Soluble Biomarkers for Better Clinical Applicability

To enhance the clinical applicability of our research, we next attempted to streamline the dataset by reducing the number of semi-quantitative ranges used to quantize biomarker values, and by considering only soluble immune mediators, *i,e.* biomarkers that can easily be quantified using ELISA-based techniques. We therefore excluded the measurement of matrix metalloproteinase (MMP)-9 activity using gelatin zymography (*Zym*) as well as all flow cytometric characterisations of immune cell subsets, methods which would be too complex for routine diagnostic application (**Supplemental Table S3**). As only cellular biomarker, we did keep the total cell count in the ‘soluble’ dataset (*TotalCellCount*) as this parameter is determined routinely in the clinic and thus readily accessible to guide treatment decisions.

Importantly, using this reduced set of biomarkers, correct classification was still possible (**Figure 5**). However, distinguishing between culture-positive infections and cases of no growth with 90%, 95% and 100% accuracy now required more individual biomarkers than before – 12, 15 and 25, respectively. Overall, our calculations demonstrated that reducing the number of semi-quantitative ranges lowered the dimensionality of the input space, enhanced clause interpretability and might simplify future biomarker tests. However, this simultaneously increased the number of biomarkers required to achieve the desired target accuracy. As such, these findings underscored the importance of cellular biomarkers that contributed key information needed for accurate classification using the smallest possible biomarker combination. **Table 3** summarises the minimal number of biomarkers for both the full original dataset versus the reduced but clinically more tractable dataset consisting of only soluble biomarkers, in relation to the numbers of semi-quantitative intervals used to Booleanise and discretise continuous biomarker values (**Figure 4; Supplemental Figures S4 and S5**).

**Figure 5.**
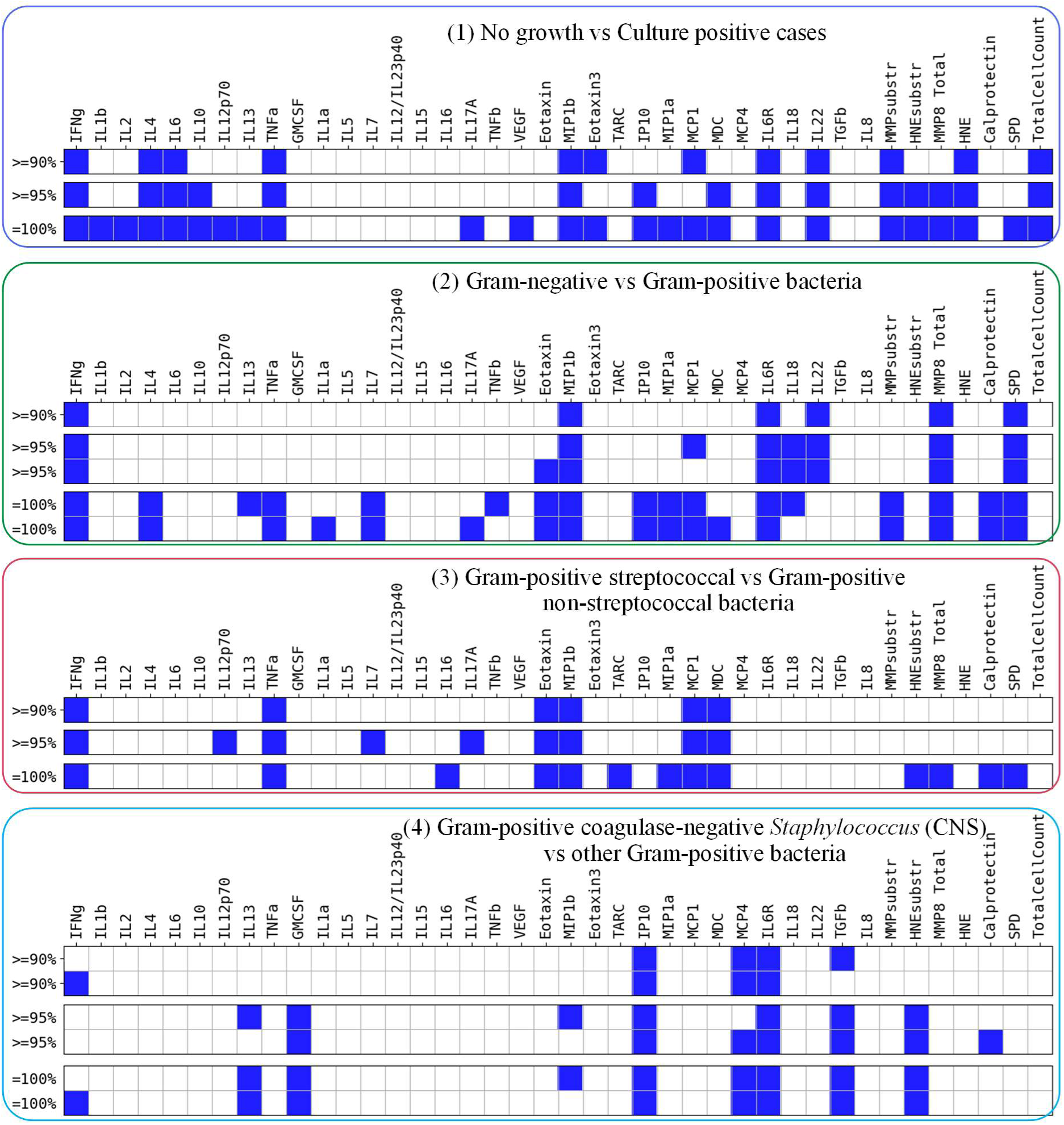
Minimised sets of soluble biomarkers (excluding *Zym* and including *TotalCellCount*) Figure shows the minimised set of soluble biomarkers needed to make predictions at different classification stages with the target accuracies of 90%, 95%, and 100% for the case where each biomarker value is represented by four semi-quantitative ranges.

**Table 3.** Minimal number of biomarkers required to achieve the target classification accuracy for different datasets and numbers of semi-quantitative ranges of biomarker values. The table reveals a trade-off between the biomarker values granularity (*i.e*. the number of semi-quantitative ranges used to quantise biomarkers) and the number of biomarkers needed to achieve the target accuracy. The exact identify of the corresponding biomarkers is shown in Figures 4 and 5 as well as in Supplemental Figures S4 and S5.

| # | Classification step | Overall classification accuracy | Full dataset<br>(40 soluble + 9 cellular biomarkers) | Soluble biomarkers<br>(excl. *Zym*, incl. *TotalCellCount*) |  |  |
| --- | --- | --- | --- | --- | --- | --- |
|  |  |  | 4 ranges | 4 ranges | 3 ranges | 2 ranges |
| 1 | No growth<br>vs Culture-positive cases | ≥ 90% | 11 (9 soluble + 3 cellular) | 12 | 12 | 19 |
|  |  | ≥ 95% | 13 (10 soluble + 3 cellular) | 15 | 16 | 24 |
|  |  | 100% | 21 (16 soluble + 5 cellular) | 25 | 33 | 33 (96.3% max. accuracy) |
| 2 | Gram-positive<br>vs Gram-negative bacteria | ≥ 90% | 6 (6 soluble) | 6 | 12 | 17 |
|  |  | ≥ 95% | 8 (6 soluble + 2 cellular) | 8 | 16 | 20 |
|  |  | 100% | 16 (11 soluble + 5 cellular) | 17 | 22 | 20 (96.8% max. accuracy) |
| 3 | Gram-positive streptococcal<br>vs non-streptococcal bacteria | ≥ 90% | 5 (3 soluble + 2 cellular) | 6 | 10 | 11 |
|  |  | ≥ 95% | 9 (7 soluble + 2 cellular) | 9 | 13 | 13 |
|  |  | 100% | 12 (9 soluble + 3 cellular) | 13 | 18 | 15 (97.8% max. accuracy) |
| 4 | Coag.-neg. *Staphylococcus*<br>vs other Gram-positive bacteria | ≥ 90% | 3 (3 soluble incl. *Zym*) | 5 | 6 | 8 |
|  |  | ≥ 95% | 6 (4 soluble + 2 cellular) | 7 | 7 | 16 |
|  |  | 100% | 7 (4 soluble + 3 cellular) | 8 | 8 | 21 |

### 3.6. Minimisation and Pruning

In addition to minimisation of the number of biomarkers used in a logical clause, the set of clauses can also be minimised, or pruned (Liu et al., 2021), based on clause efficiency and their contribution to the accurate inference. Efficiency can be estimated as a ratio between *True* and *False* predictions made by each individual clause (*i.e.* logical inference rule), which is supported by the TM and can be done by the end of training and/or testing. This ratio is shown at the top of each clause in **Figures 2 and 3**. Clauses with the minimal contribution to the true predictions (*e.g.* clauses C7 and C9 in **Figure 2**) or with a considerable portion of false predictions (*e.g.* clause C5 in **Figure 2** and clause C9 in **Figure 3**) could be pruned from the TMs. This process was repeated as long as TM performance remained higher than the target accuracy.

**Figures 6-9** present the optimised sets of TM clauses used at each classification step after minimising the number of biomarkers and removing less efficient clauses. For example, the number of soluble biomarkers was reduced from 40 to 8, followed by further pruning of the clauses from 20 per class to just 6 clauses for detecting Gram-positive bacteria and 10 clauses for identifying Gram-negative bacteria (**Figure 7**). Despite considerable minimisation, the pruned model could still successfully distinguish between Gram-positive and Gram-negative bacteria with 95% accuracy. Similar TM optimisation could be applied to all other steps of the hierarchical classification approach used in this study (**Figure 6-9**), yielding a comprehensive and overlapping set of biomarkers that defined immunologically distinct local responses during early peritonitis in patients presenting with acute symptoms (**Figure 10**).

**Figure 6.**
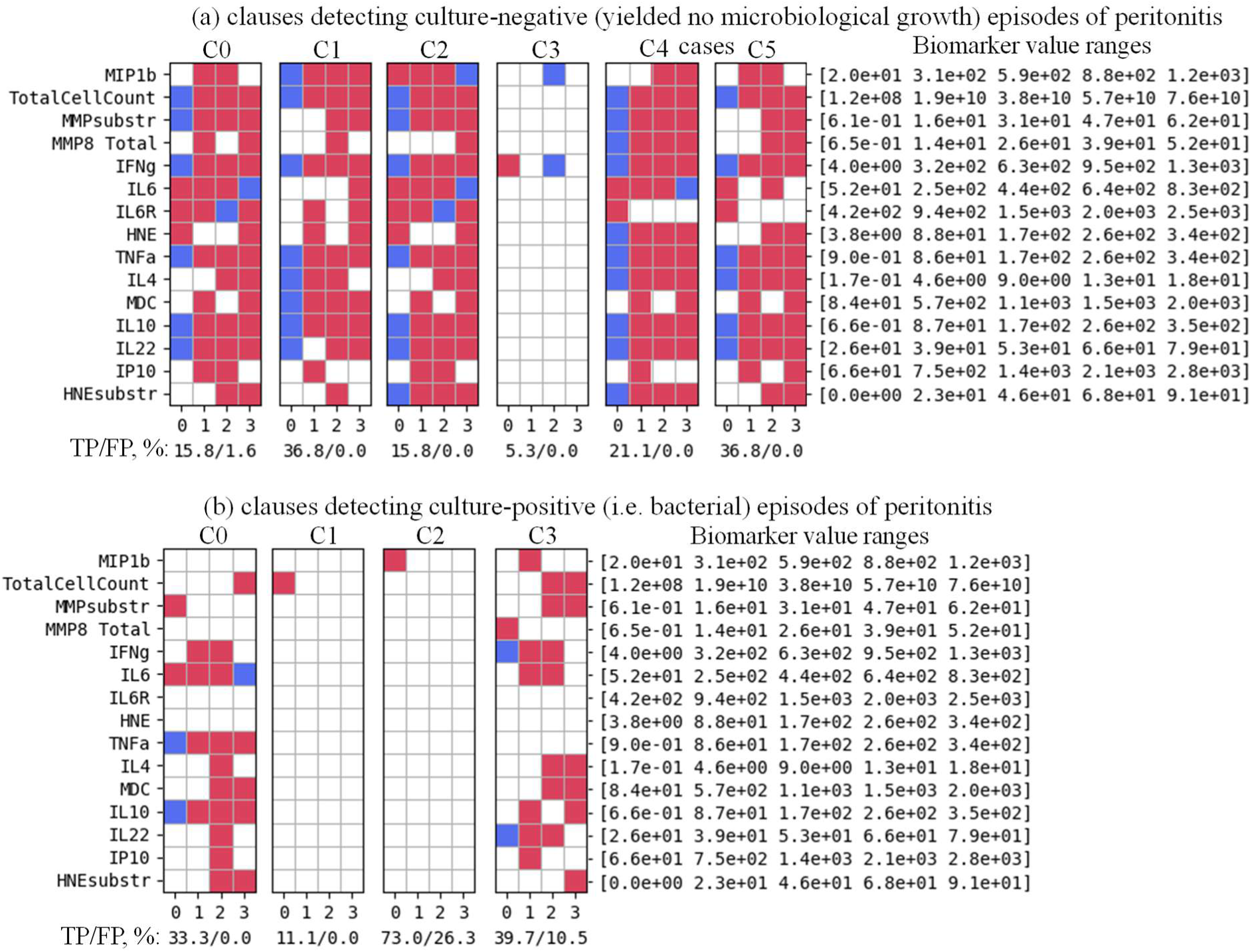
Minimised set of TM clauses and soluble biomarkers used to discriminate between episodes of peritonitis that yielded no microbiological growth versus culture-positive episodes with 95.12% accuracy for the case where each biomarker value was represented by four semi-quantitative ranges. Accuracy labels under each clause show the percentage of True Positive (TP) and False Positive (FP) predictions of individual clauses. A balance between clause generalisation and specialisation, which defines a ratio between *True* and *False* predictions made by each clause is controlled by TM hyper-parameters (Tarasyuk, Rahman, et al., 2023) and affects the overall classification accuracy.

**Figure 7.**
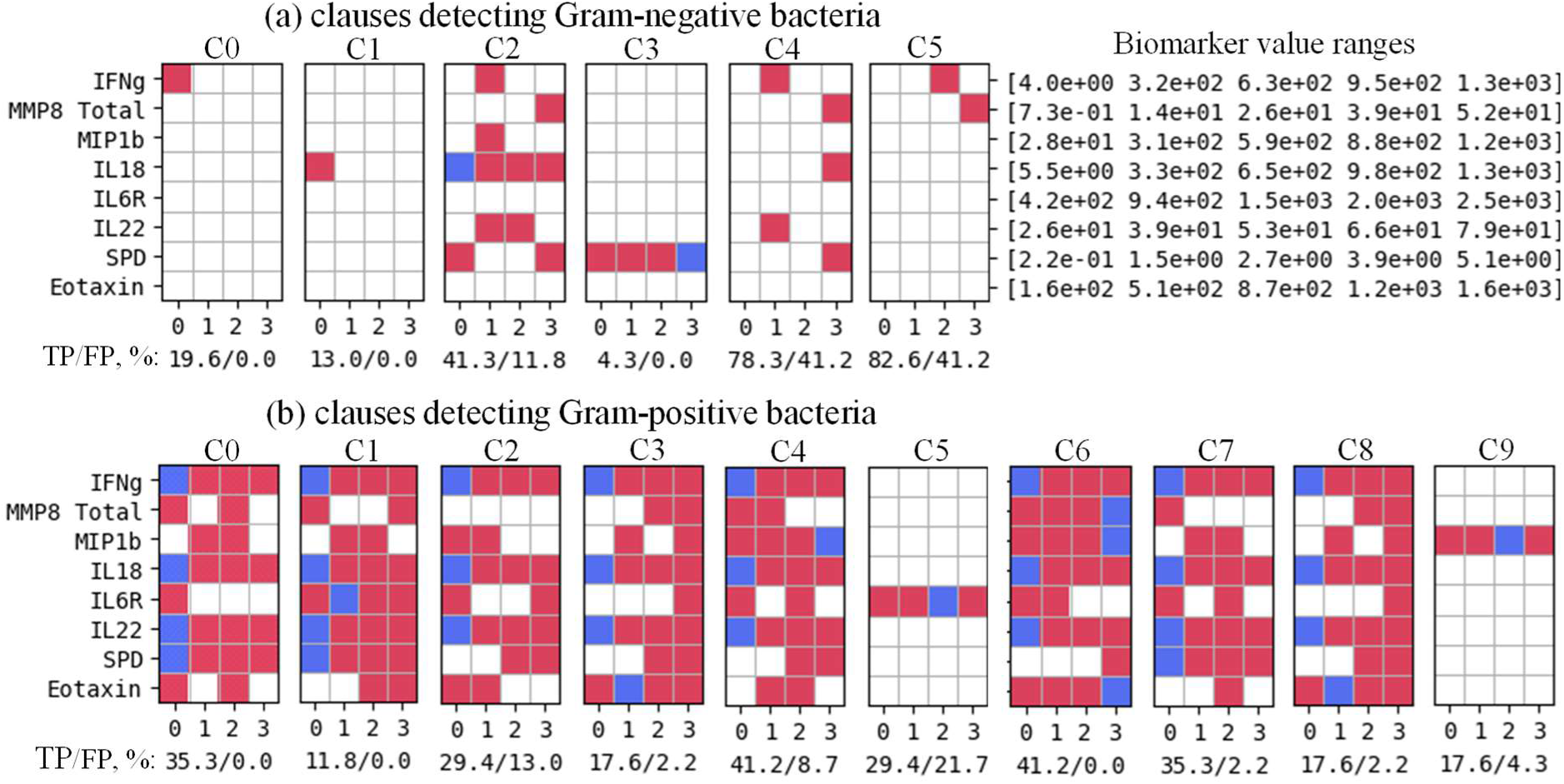
Minimised set of TM clauses and soluble biomarkers used to discriminate between episodes caused by Gram-negative bacteria within the culture-positive group of patients with 95.24% accuracy for the case where each biomarker value was represented by four semi-quantitative ranges. Accuracy labels under each clause define the percentage of True Positive (TP) and False Positive (FP) predictions of individual clauses.

**Figure 8.**
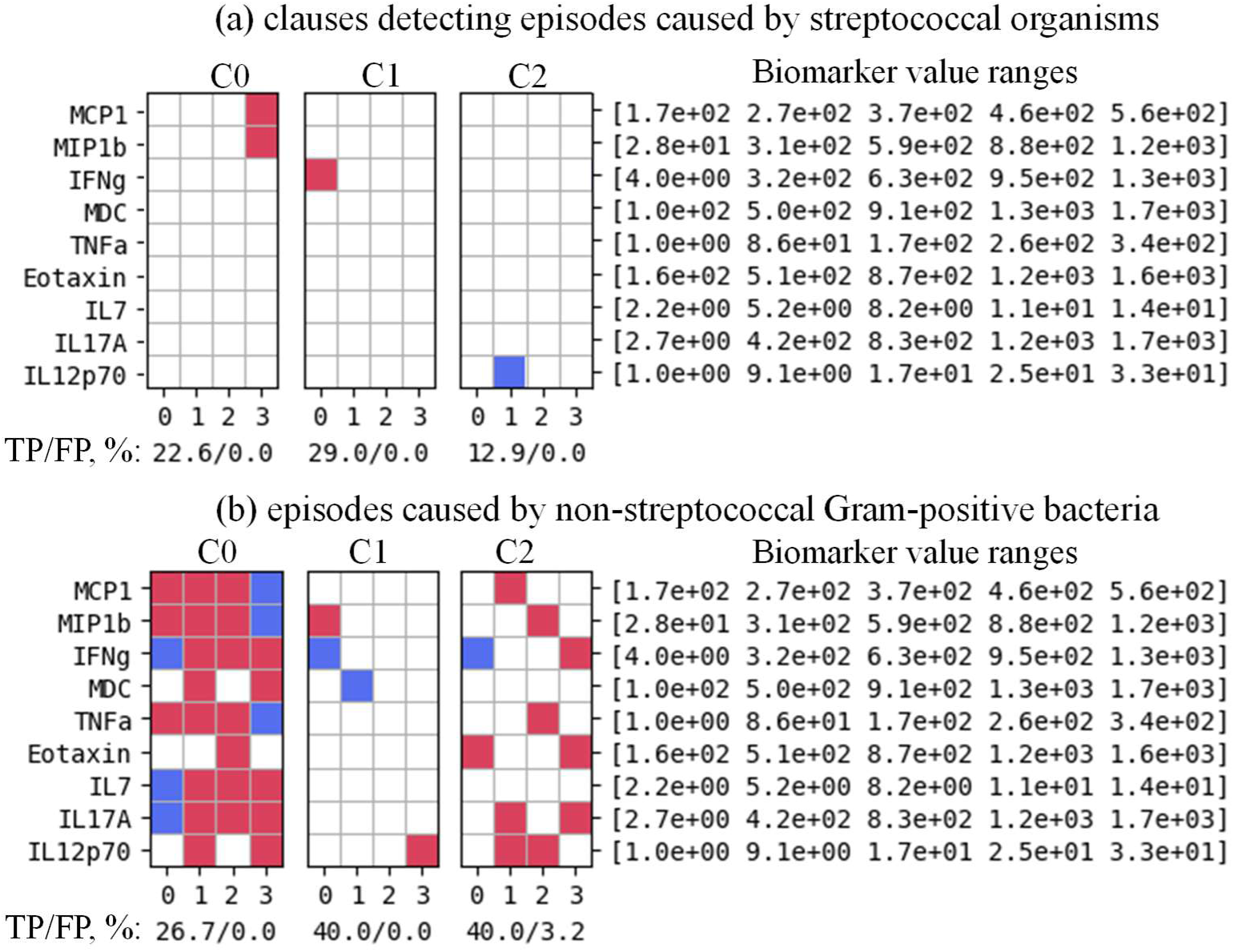
Minimised set of TM clauses and soluble biomarkers used to discriminate between episodes caused by streptococcal organisms versus episodes caused by non-streptococcal Gram-positive bacteria with 95.65% accuracy for the case where each biomarker value was represented by four semi-quantitative ranges. Accuracy labels under each clause define the percentage of True Positive (TP) and False Positive (FP) predictions of individual clauses.

**Figure 9.**
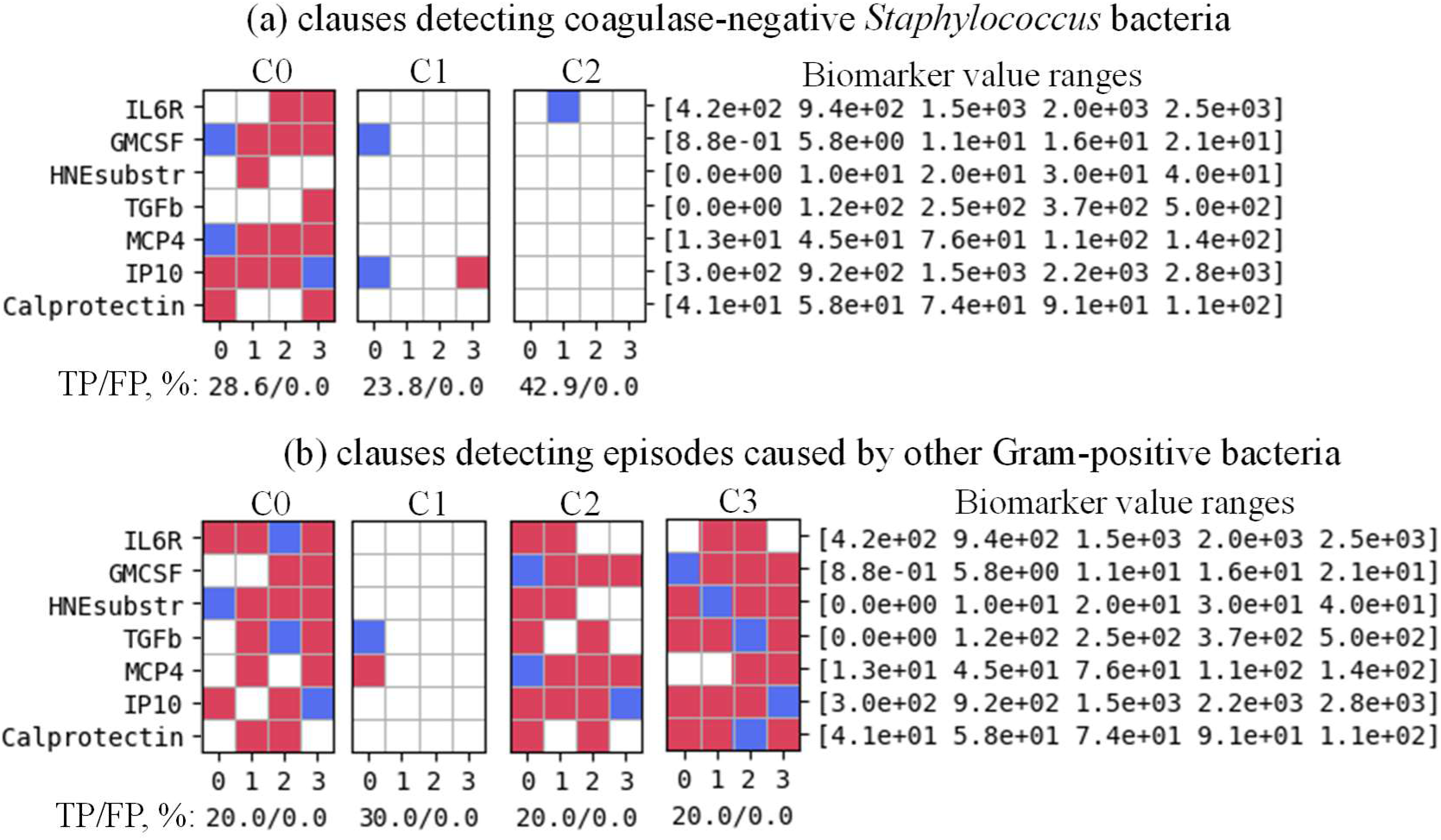
Minimised set of TM clauses and soluble biomarkers used to discriminate between episodes caused by coagulase-negative *Staphylococcus* versus episodes caused by other Gram-positive bacteria with 96.77% accuracy for the case where each biomarker value was represented by four semi-quantitative ranges. Accuracy labels under each clause define the percentage of True Positive (TP) and False Positive (FP) predictions of individual clauses.

**Figure 10.**
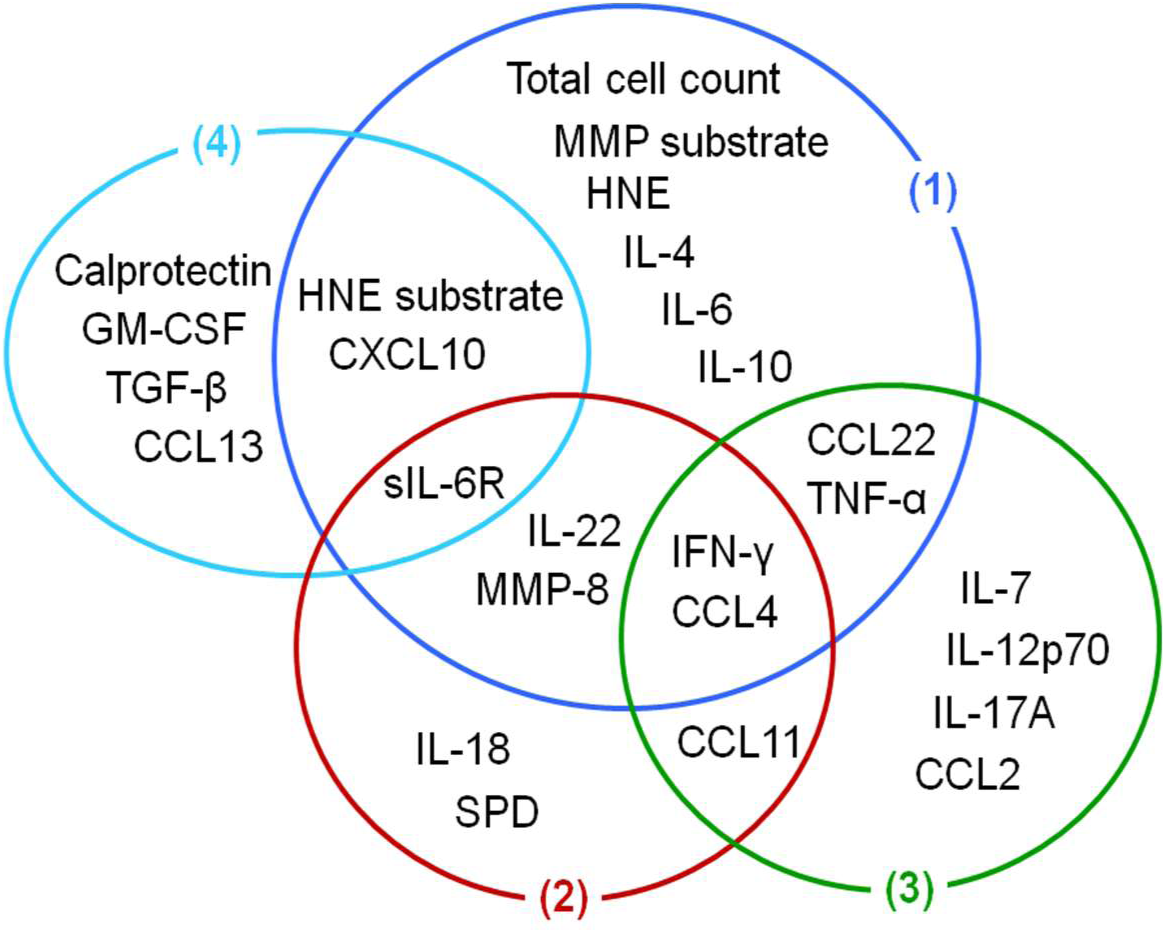
Minimised set of biomarkers defining immune fingerprints associated with peritonitis caused by different types of bacteria with an accuracy ≥95%. 1) discrimination between episodes of peritonitis that yielded no microbiological growth versus culture-positive episodes; 2) discrimination between episodes caused by Gram-negative bacteria within the culture-positive group of patients; 3) discrimination between episodes caused by streptococcal organisms versus episodes caused by non-streptococcal Gram-positive bacteria; 4) discrimination between epsodes caused by coagulase-negative *Staphylococcus* versus episodes caused by other Gram-positive bacteria.

## 4. Discussion

In this study, we demonstrate the potential of TMs as an effective ML model for analysing local immune responses in patients with life-threatening bacterial infection. By leveraging the logic-based framework of TMs, we successfully identified pathogen-specific immune fingerprints, represented as logical clauses, which are both easily interpretable and actionable for clinical decision-making. These logical rules provide insights into the distinctive biomarker profiles associated with different types of bacterial infections, enabling rapid and precise classification even before conventional microbiological results are available.

At the core, our study reaffirms the notion that different pathogens elicit qualitatively and quantitatively distinct immune responses, even when infecting the same anatomical location and causing indistinguishable clinical symptoms (Lin et al., 2013; Zhang et al., 2017). This might not come as a surprise considering that each bacterium expresses a unique set of pathogen-associated molecular patterns, antigens and virulence factors interacting with a myriad of pattern recognition factors and antigen receptors of the immune system (Kroemer et al., 2024; Medzhitov & Iwasaki, 2024). For instance, the outer membrane of Gram-negative bacteria contains lipopolysaccharides, highly immunogenic molecules that trigger inflammatory responses via Toll-like receptor 4 (TLR4) expressed on monocytes, macrophages, dendritic cell, other immune cells and many tissues. Gram-positive bacteria are free of lipopolysaccharides but can be sensed via TLR2 (Colmont et al., 2011), thus defining a clear mechanism how the body discriminates between the two main groups of bacteria. Similarly, TLR5 recognises flagellin, a principal component of flagella carried by bacteria such as *Salmonella* spp., *Pseudomonas aeruginosa*, *Listeria monocytogenes* and some strains of *E. coli* but not others (Liuzzi et al., 2015). Many microbial organisms also express the highly potent metabolite (*E*)-4-hydroxy-3-methyl-but-2-enyl pyrophosphate (HMB-PP), which specifically activates a small subset of T lymphocytes expressing a Vγ9/Vδ2 T cell receptor; notable HMB-PP deficient pathogens of clinical relevance in PD include streptococcal and staphylococcal bacteria (Liuzzi et al., 2015). Together, the unique combination of such immunogenic molecules defines each microorganism and is likely to result in immunologically distinct activation pathways.

We believe that the presence of pathogen-specific immune responses has never been documented in infected patients as clearly as in the current study, demonstrating distinct local immune responses in patients with severe infection. Of note, some of these features were particularly relevant for predicting the presence of microbiologically confirmed bacterial organisms versus cases of no growth, including the total cell count, MMP activity (*MMPsubstr*) and levels of human neutrophil elastase (HNE), IL-4, IL-6 and IL-10 in the peritoneal effluent. Others were found to play a role in predicting specific types of bacteria, such as levels of IL-18 and surfactant protein D for the discrimination between Gram-negative and Gram-positive infections; levels of IL-7, IL-12p70, IL-17A and CCL2 for the discrimination between streptococcal and non-streptococcal Gram-positive infection; and levels of calprotectin, GM-CSF, TGF-β and CCL13 for the prediction of coagulase-negative *Staphylococcus* infections versus other types of Gram-positive infections. Several biomarkers featured in immune fingerprints associated with more than one type of peritonitis, suggesting a particularly useful role for the differential diagnosis of PD patients, namely HNE activity (*HNEsubstr*) and effluent levels of IFN-γ, TNF-α, IL-22, sIL-6R, MMP-8, CCL4, CCL11, CCL22 and CXCL10. Reassuringly, despite using entirely different statistical methodologies, these patterns were remarkably similar to our earlier analyses associating TNF-α with culture-positive episodes and IL-22 and CXCL10 with Gram-positive infections (Lin et al., 2013), as well as the importance of the total cell count for culture-positive episodes, IFN-γ and IL-17A for non-streptococcal Gram-positive infections, and sIL-6R for staphylococcal infections (Zhang et al., 2017). Differences with regard to other biomarkers may in part be due to the fact that TMs work with Booleanised semi-quantitative input values rather than precise measurements as used in previous studies, and that we here used a stepwise classification of patients.

Our findings highlight the capability of TMs to address critical challenges in biomedical ML, such as interpretability and efficiency. Unlike traditional ‘black boxes’ models, TMs offer a transparent decision-making process, which is essential in clinical settings where understanding and trust in predictive models are paramount. Additionally, the ability of TMs to operate on Booleanised, semi-quantitative data underscores their suitability for mass clinical use, particularly in combination with rapid, accessible testing methods like lateral flow tests. Recent advances in lateral flow technology have already demonstrated the ability to determine multiple levels of analyte concentration (Hu et al., 2017; You et al., 2018), rather than simply detect the presence (or absence) of an individual biomarker. In this respect, a recently developed lateral flow test for diagnosis of peritonitis may not only be useful for the detection of early infection based on elevated levels of IL-6 and MMP-8 in PD effluent (Goodlad et al., 2020) but in a more quantitative way even contribute to the distinction between culture-positive episodes and cases of no growth, as suggested in the present study.

The hierarchical classification methodology employed in this study achieved high accuracy with minimal biomarker input, emphasising the strength of TMs in feature reduction and efficient data utilisation. This approach not only enhances diagnostic precision but also minimises the overall number of tests required, reducing both costs and time-to-diagnosis. In conclusion, TMs present a robust framework for decoding and visualising complex immune responses, offering a promising avenue for real-time, interpretable diagnostics in infectious disease management. Future work will focus on expanding the application of TMs to larger datasets and diverse infectious agents, potentially broadening its utility in clinical diagnostics and personalised medicine.

## Data Availability

All underlying tools for the present analysis are available at https://github.com/anatoliy-gorbenko/biomarkers-visualization. The anonymised patient data underlying this article will be shared on reasonable request to the corresponding author.

## Acknowledgements

We would like to thank all patients for participating in this study, the clinicians and nurses at the Renal Unit, University Hospital of Wales, Cardiff, for their support in sample collection, and past members and collaborators of the Eberl group for the biomarker measurements. We are especially grateful to Gloria Owens and Janet Williams in their roles as patient representatives affiliated with the Welsh Kidney Patients Association for encouraging us to pursue our research on novel ways to diagnose peritonitis in the first place.

