## Supplemental Information for "Prediction of the infecting organism in peritoneal dialysis patients with acute peritonitis using interpretable Tsetlin Machines"

**Supplemental Table S1. Basic demographics and microbiological culture results of all patient samples analysed in the present study.**

| Sample | Gender | Age range | Microbiological culture result |
|---|---|---|---|
| No growth | | | |
| NOG-01 | M | 81-85 | No growth |
| NOG-02 | M | 61-65 | No growth |
| NOG-03 | M | 61-65 | No growth |
| NOG-04 | M | 61-65 | No growth |
| NOG-05 | M | 61-65 | No growth |
| NOG-06 | M | 61-65 | No growth |
| NOG-07 | M | 61-65 | No growth |
| NOG-08 | F | 66-70 | No growth |
| NOG-09 | M | 71-75 | No growth |
| NOG-10 | F | 61-65 | No growth |
| NOG-11 | M | 41-45 | No growth |
| NOG-12 | F | 31-35 | No growth |
| NOG-13 | F | 86-90 | No growth |
| NOG-14 | F | 61-65 | No growth |
| NOG-15 | F | 61-65 | No growth |
| NOG-16 | F | 71-75 | No growth |
| NOG-17 | M | 61-65 | No growth |
| NOG-18 | M | 71-75 | No growth |
| NOG-19 | F | 51-55 | No growth |
| Gram-negative bacteria | | | |
| GN-01 | M | 76-80 | *Acinetobacter* sp. |
| GN-02 | F | 51-55 | *Acinetobacter baumannii* |
| GN-03 | M | 76-80 | *Enterobacter* sp. |
| GN-04 | M | 56-60 | *Escherichia coli* |
| GN-05 | F | 51-55 | *Escherichia coli* |
| GN-06 | F | 61-65 | *Escherichia coli* |
| GN-07 | F | 31-35 | *Escherichia coli* |
| GN-08 | M | 76-80 | *Escherichia coli* |
| GN-09 | M | 71-75 | *Escherichia coli* |
| GN-10 | F | 71-75 | *Escherichia coli* |
| GN-11 | M | 66-70 | *Enterobacter* sp. |
| GN-12 | F | 81-85 | *Morganella morganii* |
| GN-13 | F | 66-70 | *Proteus vulgaris* |
| GN-14 | M | 71-75 | *Pseudomonas aeruginosa* |
| GN-15 | M | 71-75 | *Pseudomonas aeruginosa* |
| GN-16 | F | 72.3 | *Pseudomonas aeruginosa*, anaerobic Gram-negative bacilli |
| GN-17 | M | 90.2 | Gram-negative bacilli |
| Gram-positive bacteria, streptococcal organisms | | | |
| GPS-01 | M | 71-75 | Alpha-haemolytic *Streptococcus* |
| GPS-02 | M | 61-65 | Alpha-haemolytic *Streptococcus* |
| GPS-03 | M | 81-85 | Alpha-haemolytic *Streptococcus* |

| | | | |
|---|---|---|---|
| GPS-04 | M | 76-80 | Alpha-haemolytic *Streptococcus* |
| GPS-05 | M | 66-70 | Alpha-haemolytic *Streptococcus* |
| GPS-06 | M | 66-70 | Alpha-haemolytic *Streptococcus* |
| GPS-07 | M | 86-90 | Alpha haemolytic *Streptococcus* |
| GPS-08 | M | 76-80 | Alpha-haemolytic *Streptococcus*, other *Streptococcus* sp. |
| GPS-09 | F | 61-65 | *Enterococcus faecalis* |
| GPS-10 | M | 41-45 | *Enterococcus faecium*, group A *Streptococcus* |
| GPS-11 | M | 56-60 | Microaerophilic *Streptococcus* |
| GPS-12 | M | 81-85 | Microaerophilic *Streptococcus* |
| GPS-13 | F | 56-60 | *Streptococcus* B |
| GPS-14 | F | 66-70 | *Streptococcus sanguinis* |
| GPS-15 | M | 81-85 | Vancomycin-resistant *Enterococcus* (VRE) |
| Gram-positive bacteria, CNS | | | |
| CNS-01 | M | 61-65 | Coagulase-negative *Staphylococcus* |
| CNS-02 | M | 61-65 | Coagulase-negative *Staphylococcus* |
| CNS-03 | M | 66-70 | Coagulase-negative *Staphylococcus* |
| CNS-04 | F | 36-40 | Coagulase-negative *Staphylococcus* |
| CNS-05 | F | 56-60 | Coagulase-negative *Staphylococcus* |
| CNS-06 | M | 66-70 | Coagulase-negative *Staphylococcus* |
| CNS-07 | F | 71-75 | Coagulase-negative *Staphylococcus* |
| CNS-08 | M | 51-55 | Coagulase-negative *Staphylococcus* |
| CNS-09 | F | 46-50 | Coagulase-negative *Staphylococcus* |
| CNS-10 | F | 61-65 | Coagulase-negative *Staphylococcus* |
| CNS-11 | M | 91-95 | Coagulase-negative *Staphylococcus* |
| CNS-12 | M | 31-35 | Coagulase-negative *Staphylococcus* |
| CNS-13 | M | 81-85 | Coagulase-negative *Staphylococcus* |
| CNS-14 | M | 76-80 | Coagulase-negative *Staphylococcus* |
| CNS-15 | F | 66-70 | Coagulase-negative *Staphylococcus* |
| CNS-16 | M | 41-45 | Coagulase-negative *Staphylococcus* |
| CNS-17 | M | 71-75 | Coagulase-negative *Staphylococcus* |
| CNS-18 | M | 76-80 | Coagulase-negative *Staphylococcus* |
| CNS-19 | M | 76-80 | Coagulase-negative *Staphylococcus* |
| CNS-20 | M | 76-80 | Coagulase-negative *Staphylococcus* |
| CNS-21 | F | 56-60 | Coagulase-negative *Staphylococcus* |
| Gram-positive bacteria, other | | | |
| GPO-01 | F | 66-70 | *Corynebacterium amycolatum* |
| GPO-02 | F | 46-50 | Coryneform bacteria |
| GPO-03 | M | 66-70 | Coryneform bacteria, Gram-positive bacilli |
| GPO-04 | M | 61-65 | Coryneform bacteria, coagulase-negative *Staphylococcus* |
| GPO-05 | F | 36-40 | *Staphylococcus aureus* |
| GPO-06 | M | 81-85 | *Staphylococcus aureus* |
| GPO-07 | M | 71-75 | *Staphylococcus aureus* |
| GPO-08 | M | 76-80 | *Staphylococcus aureus* |
| GPO-09 | M | 46-50 | *Staphylococcus aureus* |
| GPO-10 | F | 36-40 | *Staphylococcus aureus* (MRSA) |

**Supplemental Table S2. Demographics of patient samples analysed in the present study.** CNS, coagulase-negative *Staphylococcus*; SEM, standard error of the mean.

| | No growth | | Gram-negative | | *Streptococcus* spp. | | CNS | | Other Gram-positive | |
|---|---|---|---|---|---|---|---|---|---|---|
| | *n* | % | *n* | % | *n* | % | *n* | % | *n* | % |
| Age (mean ± SEM) | 63.8 ± 2.9 | | 68.6 ± 3.3 | | 70.6 ± 3.1 | | 65.2 ± 3.4 | | 60.9 ± 4.6 | |
| [18-40] | 1 | 5.3 | 1 | 5.9 | 0 | 0 | 2 | 9.5 | 2 | 18.2 |
| [40-50] | 1 | 5.3 | 0 | 0 | 1 | 6.7 | 1 | 4.8 | 1 | 9.1 |
| [50-60] | 1 | 5.3 | 3 | 17.6 | 2 | 13.3 | 3 | 14.3 | 1 | 9.1 |
| [60-70] | 11 | 57.9 | 3 | 17.6 | 4 | 26.7 | 6 | 28.6 | 4 | 36.4 |
| [70-80] | 3 | 15.8 | 8 | 47.1 | 3 | 20.0 | 7 | 33.3 | 2 | 18.2 |
| ≥80 | 2 | 10.5 | 2 | 11.8 | 5 | 33.3 | 2 | 9.5 | 1 | 9.1 |
| Sex | | | | | | | | | | |
| Male | 11 | 57.9 | 9 | 52.9 | 12 | 80 | 14 | 66.7 | 6 | 54.5 |
| Female | 8 | 42.1 | 8 | 47.1 | 3 | 20 | 7 | 33.3 | 5 | 45.5 |

**Supplemental Table S3. Soluble and cellular immune biomarkers in peritoneal effluent.** Methodological details were described before (Zhang et al., 2017). BD, BD Biosciences; MSD, Meso Scale Discovery.

| Biomarker | Symbol | Method, manufacturer |
|---|---|---|
| IL-1α (pg/ml) | IL1a | V-PLEX Cytokine 30-Plex Kit, MSD |
| IL-1β (pg/ml) | IL1b | V-PLEX Cytokine 30-Plex Kit, MSD |
| IL-2 (pg/ml) | IL2 | V-PLEX Cytokine 30-Plex Kit, MSD |
| IL-4 (pg/ml) | IL4 | V-PLEX Cytokine 30-Plex Kit, MSD |
| IL-5 (pg/ml) | IL5 | V-PLEX Cytokine 30-Plex Kit, MSD |
| IL-6 (pg/ml) | IL6 | V-PLEX Cytokine 30-Plex Kit, MSD |
| IL-7 (pg/ml) | IL7 | V-PLEX Cytokine 30-Plex Kit, MSD |
| IL-10 (pg/ml) | IL10 | V-PLEX Cytokine 30-Plex Kit, MSD |
| IL-12p40 (pg/ml) | ILp40 | V-PLEX Cytokine 30-Plex Kit, MSD |
| IL-12p70 (pg/ml) | ILp70 | V-PLEX Cytokine 30-Plex Kit, MSD |
| IL-13 (pg/ml) | IL13 | V-PLEX Cytokine 30-Plex Kit, MSD |
| IL-15 (pg/ml) | IL15 | V-PLEX Cytokine 30-Plex Kit, MSD |
| IL-16 (pg/ml) | IL16 | V-PLEX Cytokine 30-Plex Kit, MSD |
| IL-17A (pg/ml) | IL17A | V-PLEX Cytokine 30-Plex Kit, MSD |
| IL-18 (pg/ml) | IL18 | Single-plex assay, MSD |
| IL-22 (pg/ml) | IL22 | Single-plex assay (customised), MSD |
| sIL-6R (pg/ml) | IL6R | Single-plex assay, MSD |
| IFN-γ (pg/ml) | IFNg | V-PLEX Cytokine 30-Plex Kit, MSD |
| TNF-α (pg/ml) | TNFa | V-PLEX Cytokine 30-Plex Kit, MSD |
| TNF-β (pg/ml) | TNFb | V-PLEX Cytokine 30-Plex Kit, MSD |
| GM-CSF (pg/ml) | GMCSF | V-PLEX Cytokine 30-Plex Kit, MSD |
| TGF-β (pg/ml) | TGFb | ELISA, R&D Systems |
| VEGF (pg/ml) | VEGF | V-PLEX Cytokine 30-Plex Kit, MSD |
| CCL2 (pg/ml) | MCP1 | ELISA, BD Biosciences |
| CCL3 (pg/ml) | MIP1a | V-PLEX Cytokine 30-Plex Kit, MSD |
| CCL4 (pg/ml) | MIP1b | V-PLEX Cytokine 30-Plex Kit, MSD |
| CCL11 (pg/ml) | Eotaxin | V-PLEX Cytokine 30-Plex Kit, MSD |
| CCL13 (pg/ml) | MCP4 | V-PLEX Cytokine 30-Plex Kit, MSD |
| CCL17 (pg/ml) | TARC | V-PLEX Cytokine 30-Plex Kit, MSD |
| CCL22 (pg/ml) | MDC | V-PLEX Cytokine 30-Plex Kit, MSD |
| CCL26 (pg/ml) | Eotaxin3 | V-PLEX Cytokine 30-Plex Kit, MSD |
| CXCL8 (pg/ml) | IL8 | V-PLEX Cytokine 30-Plex Kit, MSD |
| CXCL10 (pg/ml) | IP10 | V-PLEX Cytokine 30-Plex Kit, MSD |
| MMP-8 total (ng/ml) | MMP8 Total | ELISA, R&D Systems DuoSet |
| MMP-9 activity (arbitrary units) | Zym | Gelatin zymography, Invitrogen NuPage |
| MMP substrate (ng/ml) | MMPsubstr | Enzo Life Sciences |
| Human neutrophil elastase (ng/ml) | HNE | ELISA, Mologic |
| HNE substrate (ng/ml) | HNEsubstr | Bachem |
| Calprotectin (ng/ml) | Calprotectin | ELISA, Hycult |
| Surfactant protein D (ng/ml) | SPD | ELISA, R&D Systems DuoSet |

| | | |
|---|---|---|
| Total cell count (× $10^9$ cells) | TotalCellCount | Microscopy |
| $CD3^+$ (% of total) | CD3+live(%) | Flow cytometry, BD FACSCanto II |
| $CD14^+$ (% of total) | CD14+live(%) | Flow cytometry, BD FACSCanto II |
| $CD15^+$ (% of total) | CD15+live(%) | Flow cytometry, BD FACSCanto II |
| CD4:CD8 ratio | CD4/CD8 | Flow cytometry, BD FACSCanto II |
| $CD4^+$ (% of T cells) | CD4/CD3(%) | Flow cytometry, BD FACSCanto II |
| $CD8^+$ (% of T cells) | CD8/CD3(%) | Flow cytometry, BD FACSCanto II |
| $V\gamma9^+$ (% of T cells) | g9T/CD3(%) | Flow cytometry, BD FACSCanto II |
| $V\delta2^+$ (% of T cells) | d2T/CD3(%) | Flow cytometry, BD FACSCanto II |

**Supplemental Table S4. Proportion of missing values imputed using Multivariate Imputation by Chained Equations (MICE).**

| **Biomarker** | **Symbol** | **% of missing values** |
|---|---|---|
| IL-18 (pg/ml) | `IL18` | 3.6 |
| Total cell count (× $10^9$ cells) | `TotalCellCount` | 4.8 |
| IL-22 (pg/ml) | `IL22` | 6.0 |
| MMP substrate (ng/ml) | `MMPsubstr` | 6.0 |
| HNE substrate (ng/ml) | `HNEsubstr` | 6.0 |
| MMP-9 activity (arbitrary units) | `Zym` | 6.0 |
| MMP-8 total (ng/ml) | `MMP8 Total` | 6.0 |
| Human neutrophil elastase (ng/ml) | `HNE` | 6.0 |
| Calprotectin (ng/ml) | `Calprotectin` | 6.0 |
| Surfactant protein D (ng/ml) | `SPD` | 6.0 |
| $CD3^+$ (% of total) | `CD3+live(%)` | 12.1 |
| $CD14^+$ (% of total) | `CD14+live(%)` | 12.1 |
| $CD15^+$ (% of total) | `CD15+live(%)` | 12.1 |
| $V\gamma 9^+$ (% of T cells) | `g9T/CD3(%)` | 12.1 |
| CD4:CD8 ratio | `CD4/CD8` | 16.9 |
| $CD4^+$ (% of T cells) | `CD4/CD3(%)` | 16.9 |
| $CD8^+$ (% of T cells) | `CD8/CD3(%)` | 16.9 |
| $V\delta 2^+$ (% of T cells) | `d2T/CD3(%)` | 16.9 |

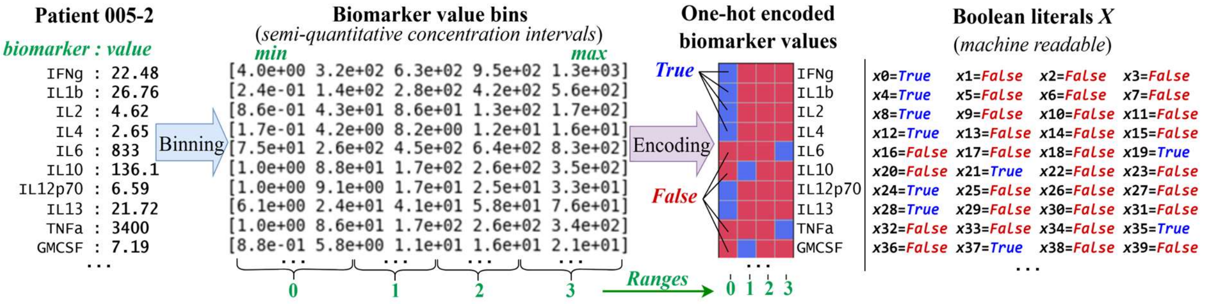


**Supplemental Figure S1. Data Booleanisation: binning, encoding and visualisation.** The figure shows sample #005-2 as example, from an individual infected with *Proteus vulgaris* (a Gram-negative bacterium). As a result of the Booleanisation, each biomarker value was replaced by a unique binary vector of Boolean features $X=\{x_i\}$ with the same length as the number of used semi-quantitative intervals. In this vector, only one element was set to 1 (*i.e. True;* blue pixel), indicating the presence of the biomarker value in that specific interval, and all other elements were set to 0 (*i.e. False*; red pixel).

```
Clause #0:  x0 ∧ ¬x1 ∧ ¬x2 ∧ ¬x3 ∧ ¬x6 ∧ ¬x7 ∧  x8 ∧ ¬x9 ∧ ¬x10 ∧ ¬x11 ∧ …
Clause #1:  x0 ∧ ¬x1 ∧ ¬x2 ∧ ¬x3 ∧  x4 ∧ ¬x5 ∧ ¬x6 ∧ ¬x7 ∧  x8  ∧ ¬x9  ∧ …
Clause #2:  x0 ∧ ¬x1 ∧ ¬x2 ∧ ¬x3 ∧  x4 ∧ ¬x5 ∧ ¬x6 ∧ ¬x7 ∧  x8  ∧ ¬x9  ∧ …
Clause #3:  x0 ∧ ¬x1 ∧ ¬x2 ∧ ¬x3 ∧ ¬x6 ∧ ¬x7 ∧  x8 ∧ ¬x9 ∧ ¬x10 ∧ ¬x11 ∧ …
Clause #4:  x0 ∧ ¬x1 ∧ ¬x2 ∧ ¬x3 ∧  x4 ∧ ¬x5 ∧ ¬x6 ∧ ¬x7 ∧  x8  ∧ ¬x9  ∧ …
Clause #5: ¬x112 ∧ x114 ∧ ¬x183  ∧ x185 ∧ ¬x192
Clause #6: ¬x21  ∧ ¬x42 ∧ ¬x91   ∧ ¬x117 ∧  x147 ∧ ¬x148   ∧ x157
Clause #7:  x0 ∧ ¬x1 ∧ ¬x2 ∧ ¬x3 ∧  x4   ∧ ¬x5 ∧ ¬x6 ∧ ¬x7 ∧ x8 ∧ ¬x9  ∧ …
Clause #8:  x82  ∧ x114 ∧ x144   ∧ x170
Clause #9: ¬x12  ∧ x156
```

**Supplemental Figure S2. Positive clauses (logical rules) recognising Gram-negative bacterial infection.** Each clause specifies which Boolean features $X = \{x_i\}$ of a patient sample (see Figure 2) must be *True*, *False* or should be ignored (if the feature is not included in the clause) to support the decision that the patient is infected with Gram-negative bacteria. These logical rules corresponded to clauses inference visualised in Figure 3.

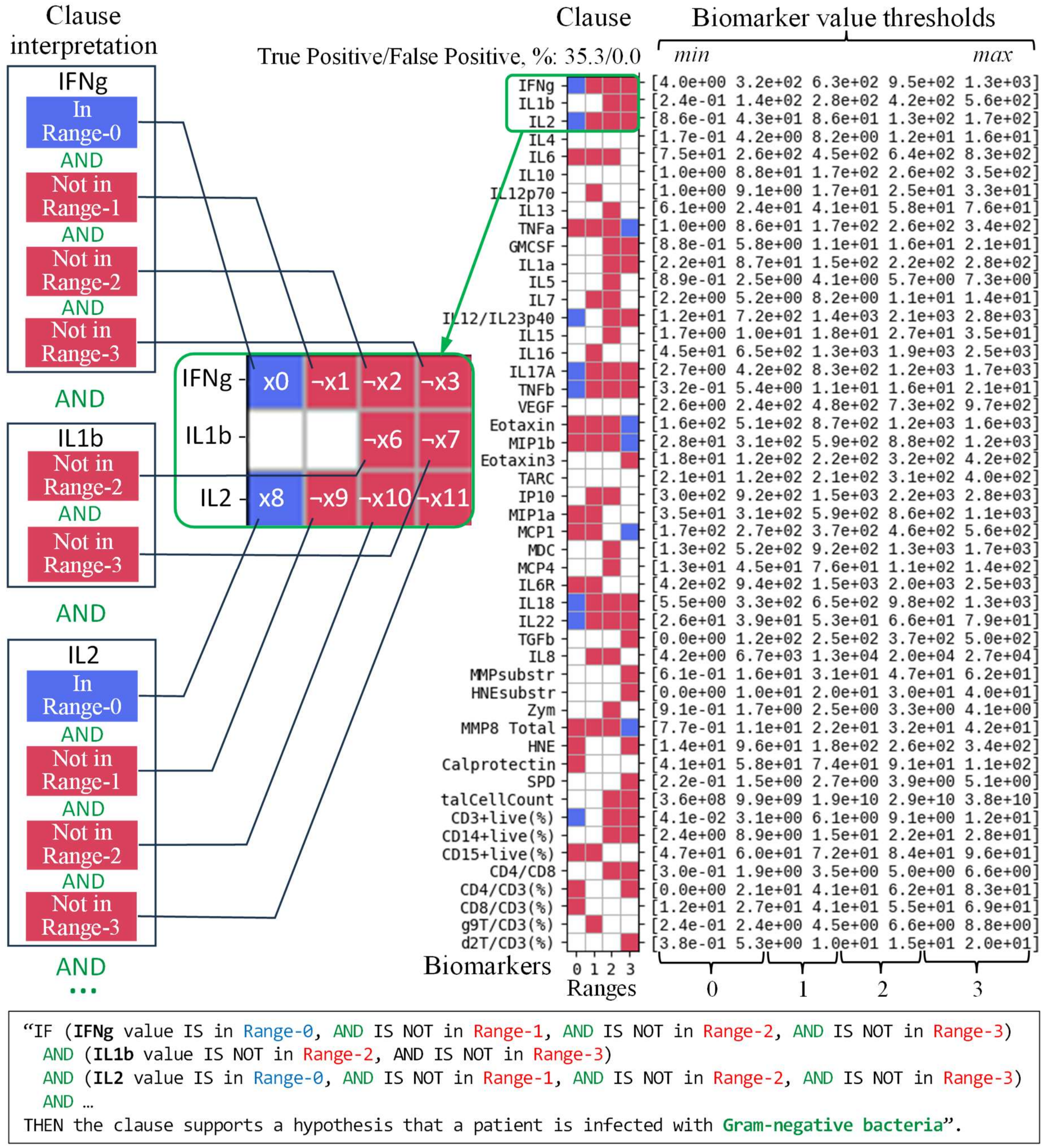


**Supplemental Figure S3. Clause visualisation and interpretation.** The figure presents an example of a clause supporting decision-making in favour of Gram-negative bacterial infection. A blue pixel means that the corresponding Boolean feature must be *True* (*i.e.* the biomarker value must be within that specific range) for a patient sample to be classified as Gram-negative bacterial infection. Red means that that the corresponding Boolean feature must be *False* (*i.e.* the biomarker value must not be in that range). Finally, white means that the corresponding Boolean feature was excluded from the clause (*i.e.* the biomarker value may or may not be in that range as it does not affect the classification).

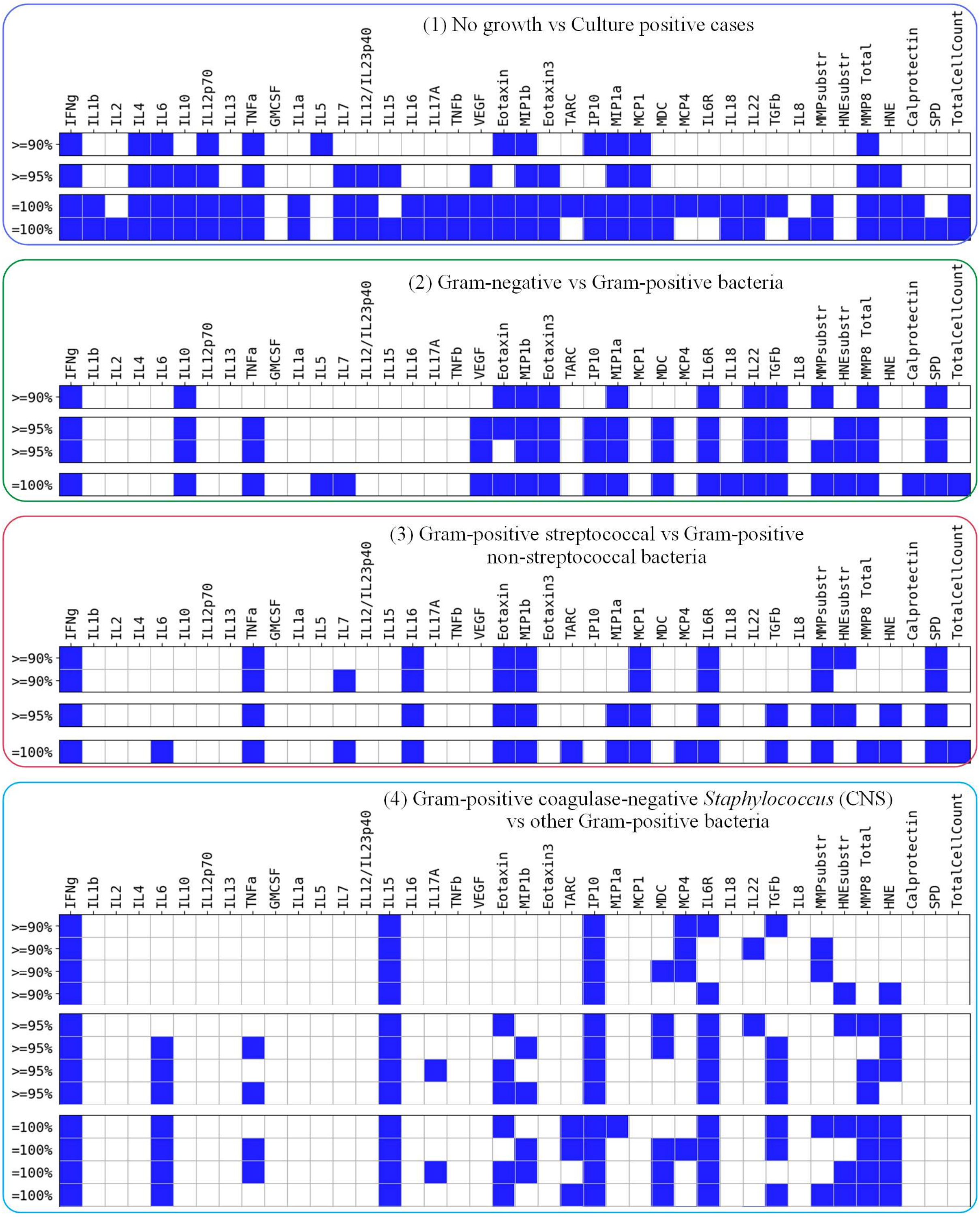


**Supplemental Figure S4. Minimised sets of soluble immune biomarkers for the case where each biomarker value was Booleanised by three semi-quantitative ranges.** Figure shows the minimised set of soluble biomarkers (excluding *Zym*, including *TotalCellCount*) needed to make predictions at different classification stages with the target accuracies of 90%, 95% and 100%, respectively.

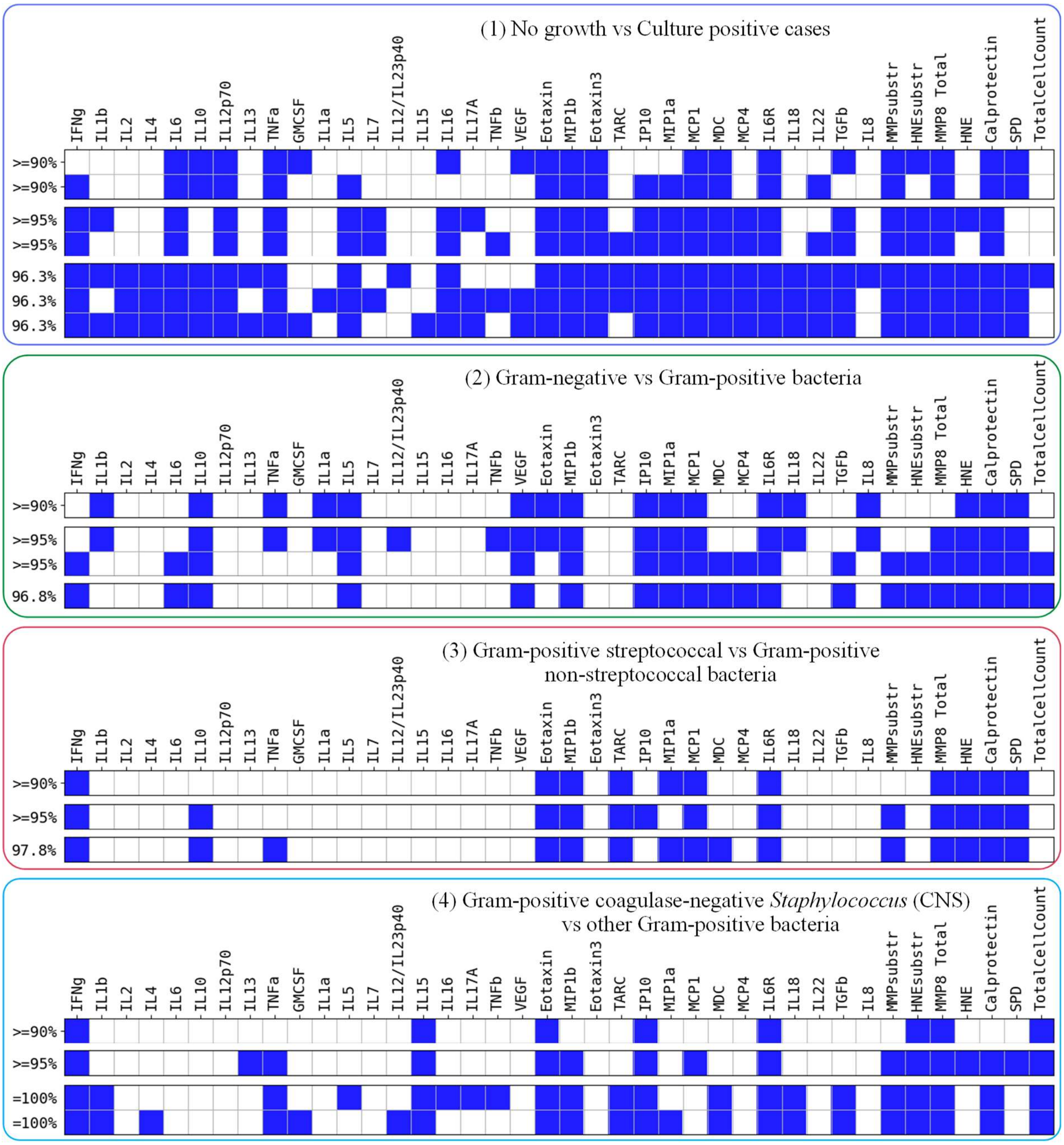


**Supplemental Figure S5. Minimised sets of soluble immune biomarkers for the case where each biomarker value was Booleanised by two semi-quantitative ranges.** Figure shows the minimised set of soluble biomarkers (excluding *Zym*, including *TotalCellCount*) needed to make predictions at different classification stages with the target accuracies of 90%, 95% and 100%, respectively.
